# Promoting inclusivity and acceptance of diverse sexual and gender identities in schools: a Rapid Realist Review of universal interventions to improve mental health

**DOI:** 10.1101/2022.08.02.22277994

**Authors:** Merle Schlief, Theodora Stefanidou, Talen Wright, Grace Levy, Alexandra Pitman, Gemma Lewis

## Abstract

**Background:** Sexual minority and trans young people are more likely to experience depression, anxiety, self-harm and suicidality than their heterosexual or cisgender peers. Improving inclusivity and acceptance of diverse sexual and gender identities, through universal interventions in schools, could prevent these mental health problems. We reviewed evidence, and developed a conceptual framework, to explain which universal interventions work, for whom, in which contexts, and why.

**Methods:** We conducted a Rapid Realist Review, with a systematic search of published, peer-reviewed and grey literature. We included reports from a call for evidence and website searches. Data were extracted in Context-Mechanism-Outcome (CMO) configurations. CMOs were developed and refined through discussions with experts by lived experience including young people, teachers, school governors and policy representatives

**Findings:** We included 53 studies, and classified interventions into five themes: Gay-Straight Alliances or similar student clubs (e.g. pride clubs), inclusive anti-bullying and harassment policies, inclusive curricula, workshops, and staff training. These interventions have the potential to reduce mental health problems among sexual minority and trans young people by reducing discrimination, bullying and feelings of unsafety, exclusion and marginalisation. The interventions appear to work best when teaching staff and school leaders are properly trained, and the school climate and community are supportive. Interventions may be less effective for boys and trans and bisexual students.

**Interpretation:** Our findings provide guiding principles for schools to develop interventions to improve the mental health of sexual minority and trans students. These findings should encourage primary research to confirm, refute or refine our programme theories.

## Introduction

Depression and anxiety are the two most common mental health problems, and they often begin during adolescence.^1,2^ Self-harm is frequently co-morbid with adolescent depression and anxiety. These mental health problems are leading risk factors for suicide and suicide attempts.^3,4^ There is evidence that rates of depression, anxiety, self-harm and suicide are rising among young people.^1,5,6^ Public health interventions to prevent these mental health problems would reduce their rising incidence and alleviate the burden on clinical services.

Sexual minority young people (e.g. lesbian, gay, bisexual, queer) are twice as likely to experience depression, anxiety, self-harm and suicidality than their heterosexual peers.^7–9^ There are few high-quality population-based studies of mental health among trans (transgender, non-binary, gender diverse) compared with cisgender young people.^10,11^ However, there is evidence that trans young people are at increased risk of depression, anxiety, self-harm and suicidality.^12,13^ Around 10% of young people identify as sexual minorities.^14,15^ The proportion of young people who are trans is unclear, but estimated at around 2%.^16^

Universal interventions aim to reduce exposure to modifiable causal risk factors and have succeeded at preventing physical health problems such as heart disease and certain cancers.^17,18^ Universal interventions could transform the prevention of mental health problems,^17^ but their development continues to lag behind physical health. Schools are a potential setting for preventative interventions that would reach most young people. There is evidence that, in schools, sexual minority and trans young people experience higher levels of bullying, discrimination, exclusion and marginalisation than their heterosexual or cisgender peers.^7,19–22^ This supports minority stress theory; that the primary cause of mental health problems among sexual and gender minorities is exposure to stigma, prejudice and discrimination, in a society that promotes being heterosexual and cisgender as normal.^23,24^ Consistent with minority stress theory, universal interventions which promote inclusivity and acceptance of diverse sexual and gender identities in schools, could prevent or reduce mental health problems among sexual minority and trans young people. This could occur through reductions in potential causal risk factors such as homophobic, biphobic and transphobic bullying, discrimination, and exclusion.

To our knowledge, no study has synthesised evidence on universal school-based interventions to promote inclusivity and acceptance of diverse sexual and gender identities. Realist approaches to evidence synthesis can be seen as complementary to systematic reviews and meta-analyses.^25^ The realist approach argues that we should do more than investigate the effectiveness of an intervention.^26^ Realistic approaches ask ‘what works for whom, in what contexts, to what extent, how and why?’ using a context-mechanism-outcome (CMO) approach. The aim is to develop, refine and test theories about how interventions trigger mechanisms, which interact with contexts, to generate outcomes.^27,28^ The context (C) refers to characteristics of the people and environments in which the intervention operates. Mechanisms (M) are the responses (e.g. internal psychosocial reactions and reasonings) triggered by changes in context (C), to generate outcomes (O). Realist syntheses use Context-Mechanism-Outcome (CMO) combinations to generate programme theories, which suggest that certain interventions are more or less likely to work, for certain people, in certain situations.^27^ We conducted a Rapid Realist Review (a time-sensitive realist synthesis),^27^ to answer the following questions:

- What universal school-based interventions to promote inclusivity and acceptance of diverse sexual and gender identities exist, and how and where were they implemented?
- In which contexts, and for whom, do these interventions work (or not work), and why?

## Methods

We used the steps outlined by Saul and colleagues:^27^

1. Developed the scope by clarifying the content area
2. Defined the research questions and ensured there was enough evidence to answer them
3. Identified how findings and recommendations would be used
4. Developed search terms and inclusion/exclusion criteria
5. Identified and screened peer-reviewed papers and data from other sources including websites and grey literature
6. Extracted and synthesised data
7. Validated findings with experts by lived experience (see below) to draw inferences and make hypotheses.

We pre-registered our protocol with the prospective register of systematic reviews, PROSPERO: https://www.crd.york.ac.uk/prospero/display_record.php?RecordID=279193.

### Consultation with experts by lived experience

#### Young Person’s Advisory Group (YPAG)

We recruited a YPAG, which consisted of eight sexual minority or trans young people (aged 14 to 24 years) with experience of depression, anxiety, self-harm or suicidality. Young people were recruited through the McPin Foundation, a leading charity placing lived experience at the heart of mental health research. One YPAG member joined our research team and worked with us on the literature search, data extraction and synthesis. We held three 1.5-hour long involvement meetings. Meeting one focused on steps 1 and 2 of the Rapid Realist Review (above). Meeting two focused on interpretating preliminary findings and how they could be used in practice (steps 3 and 7). Meeting three focused on validating findings to refine the programme theory (step 7).

#### Stakeholder Advisory Group (SAG)

We also worked with a Stakeholder Advisory Group (SAG) comprising a secondary school governor, a secondary school teacher, and two members of the UK government Department for Education (DfE). The SAG advised on what is currently happening in schools, and what would be useful and feasible from the perspective of school leaders and policy makers. We held two meetings to work on steps 1 and 2, and validated findings via email to refine the programme theory (steps 3 and 7).

### Search strategy

We searched PubMed, PsycINFO and Web of Science (search terms in the Appendix). We also consulted experts, including young people and relevant organisations, to identify grey literature. A Call for Evidence was disseminated via Twitter to invite schools, organisations, and young people to submit evidence.

### Inclusion and exclusion criteria

We included any study design (e.g. quantitative, qualitative, implementation and intervention studies). We also searched for non-peer reviewed reports posted on websites of relevant Lesbian, Gay, Bisexual, Trans and Queer (LGBTQ+) organisations. There were no restrictions in publication dates but only studies in English were included. We excluded studies that did not provide enough detail to contribute to the development of our programme theories.

#### Participants/population

We included studies of sexual minority, trans, heterosexual and cisgender students who were aged 11 to 18 years and attending secondary school. We also included studies of secondary school teaching staff. We were primarily interested in universal interventions aimed at all students and teaching staff. We included interventions aimed solely at students or staff. If a study included students under the age of 11 or above the age of 18, we reviewed its contribution to the programme theory to determine inclusion.

#### Main outcome(s)

We included the following mental health outcomes: depression, anxiety, self-harm and suicidality. We also included the following measures of inclusivity and acceptance as outcomes (even if the study did not assess mental health): bullying, school climate, school connectedness, stigma, prejudice and discrimination. This was consistent with our hypothesis that reducing these potential causal risk factors would prevent depression, anxiety, self-harm and suicidality.

### Study selection

We imported all records into the review software Rayyan and removed duplicates. Titles and abstracts were split and screened by two researchers (MS and TS). A 10% random sample was reviewed independently by a third researcher (TW). Full-texts were split and screened by five researchers. A 10% random sample was reviewed independently by a third researcher (AP). Disagreements were resolved by consensus or after discussion with the lead researcher (GL). Reasons for exclusion were recorded, acknowledging that some records might have multiple reasons for exclusion.

#### Data extraction

We used a data extraction schedule to extract: study aim(s) and design, intervention type, sample characteristics and size, and CMOs.

#### Data synthesis

We synthesized CMOs from individual studies to create overarching CMOs and develop a programme theory. Consultations with the YPAG and Stakeholder Advisory Groups prioritised our findings and refined and highlighted gaps in the programme theory. For example, both groups highlighted which findings resonated with their lived experience or work in schools or policy. They also provided feedback on the feasibility, implementation and likely effectiveness of interventions in schools.

### Quality assessment

Realist Review methodology does not usually recommend a quality assessment and focuses instead on the relevance of studies to the programme theory.^27^ During the extraction phase, we assessed each study in terms of whether the evidence contributed to theory development, and excluded studies where we could not extract CMOs.

### Data availability statement

The data that support the findings of this study are openly available in the individual studies which constitute the review. Data from the synthesis for this study are available from the corresponding author upon reasonable request.

## Results

We identified 5155 records from database searches, and 16 through other sources (Figure 1). We screened 407 full-texts and included 53 eligible studies (Figure 1): 52 peer-reviewed articles and through the call for evidence.^29^ Studies were published between 1995 and 2021, and 65% were conducted in North America (Table 1). Thirteen included data on mental health outcomes (Table 2). Full characteristics of the studies included are presented in Supplementary Table 2.

**Figure 1.**
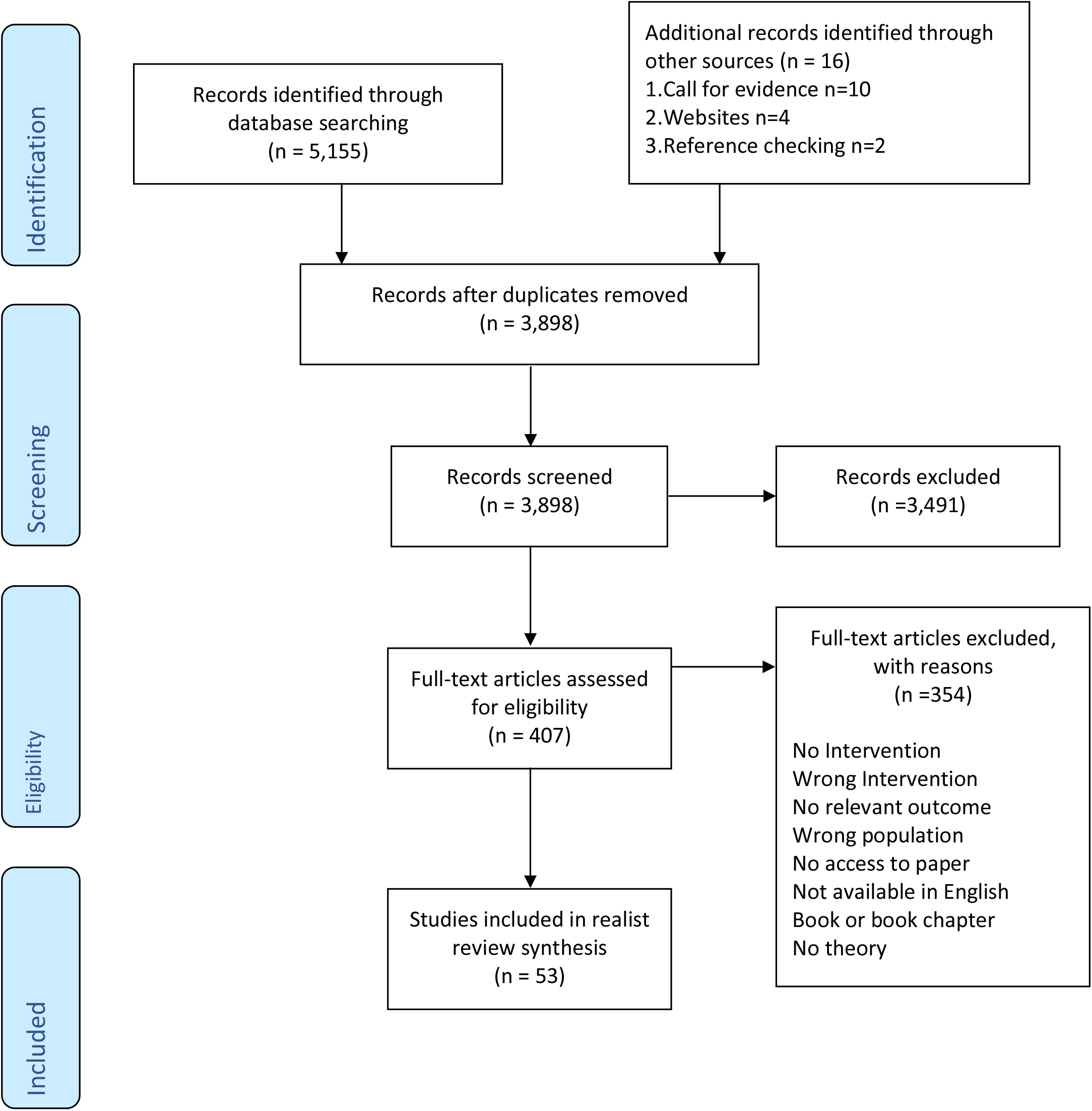
PRISMA 2009 Flow Diagram. *From:* Moher D, Liberati A, Tetzlaff J, Altman DG, The PRISMA Group (2009). *P*referred *R*eporting *I*tems for *S*ystematic Reviews and *M*eta-*A*nalyses: The PRISMA Statement. PLoS Med 6(6): e1000097. For more information, visit **www.prisma-statement.org**

**Table 1.**
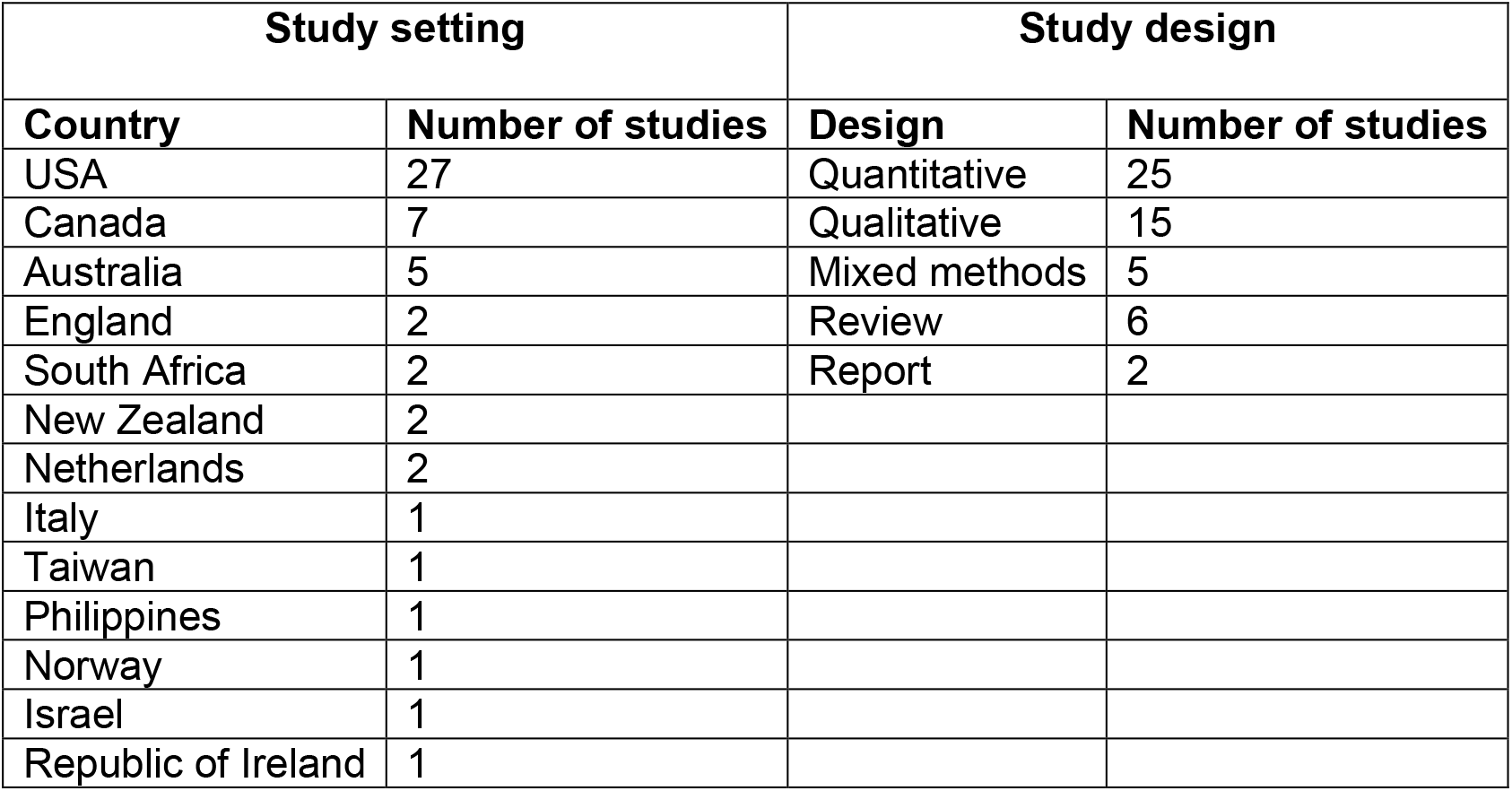
Summary of studies (n=53)

| Study setting |  | Study design |  |
| --- | --- | --- | --- |
| Country | Number of studies | Design | Number of studies |
| USA | 27 | Quantitative | 25 |
| Canada | 7 | Qualitative | 15 |
| Australia | 5 | Mixed methods | 5 |
| England | 2 | Review | 6 |
| South Africa | 2 | Report | 2 |
| New Zealand | 2 |  |  |
| Netherlands | 2 |  |  |
| Italy | 1 |  |  |
| Taiwan | 1 |  |  |
| Philippines | 1 |  |  |
| Norway | 1 |  |  |
| Israel | 1 |  |  |
| Republic of Ireland | 1 |  |  |

**Table 2.** Studies with data on mental health outcomes

| Reference | Type of intervention | Mental health outcomes |
| --- | --- | --- |
| Black et al. 2012 | GSAs and similar student clubs; inclusive anti-bullying or anti-harassment policies | Suicide attempts |
| Burk et al. 2018 | Inclusive curricula | Suicidal ideation |
| Evans and Rawlings 2021 | LGBTQ+ all staff training; inclusive curricula | General mental health and wellbeing discussed in qualitative interviews |
| Fleshman 2019 | Inclusive curricula | Mental health and wellbeing (depressed mood and suicidality) |
| Harris et al. 2021 | GSAs and similar student clubs | General mental health and wellbeing discussed in qualitative interviews |
| Hatzenbuehler and Keyes 2013 | Inclusive anti-bullying and harassment policies | Suicide attempts |
| Ioverno et al. 2016 | GSAs and similar student clubs | Wellbeing, self-esteem, and depression |
| Jones and Hillier 2012 | Inclusive anti-bullying and harassment policies | Self-harm, suicidal ideation, suicidal thoughts, and suicide attempts |
| Kosciw et al. 2012 | Inclusive curricula; Inclusive anti-bullying and harassment policies | Self-esteem |
| O'Farrell et al. 2021 | Inclusive curricula | Mental health and wellbeing |
| Saewyc et al. 2014 | GSAs and similar student clubs; Inclusive anti-bullying and harassment policies | Suicidal thoughts and suicide attempts |
| Saewyc et al. 2016 | GSAs and similar student clubs; Inclusive anti-bullying and harassment policies | Suicidal ideation |

We classified interventions into five themes (full details of the interventions and what they involve are provided in Table 3):

**Table 3.**
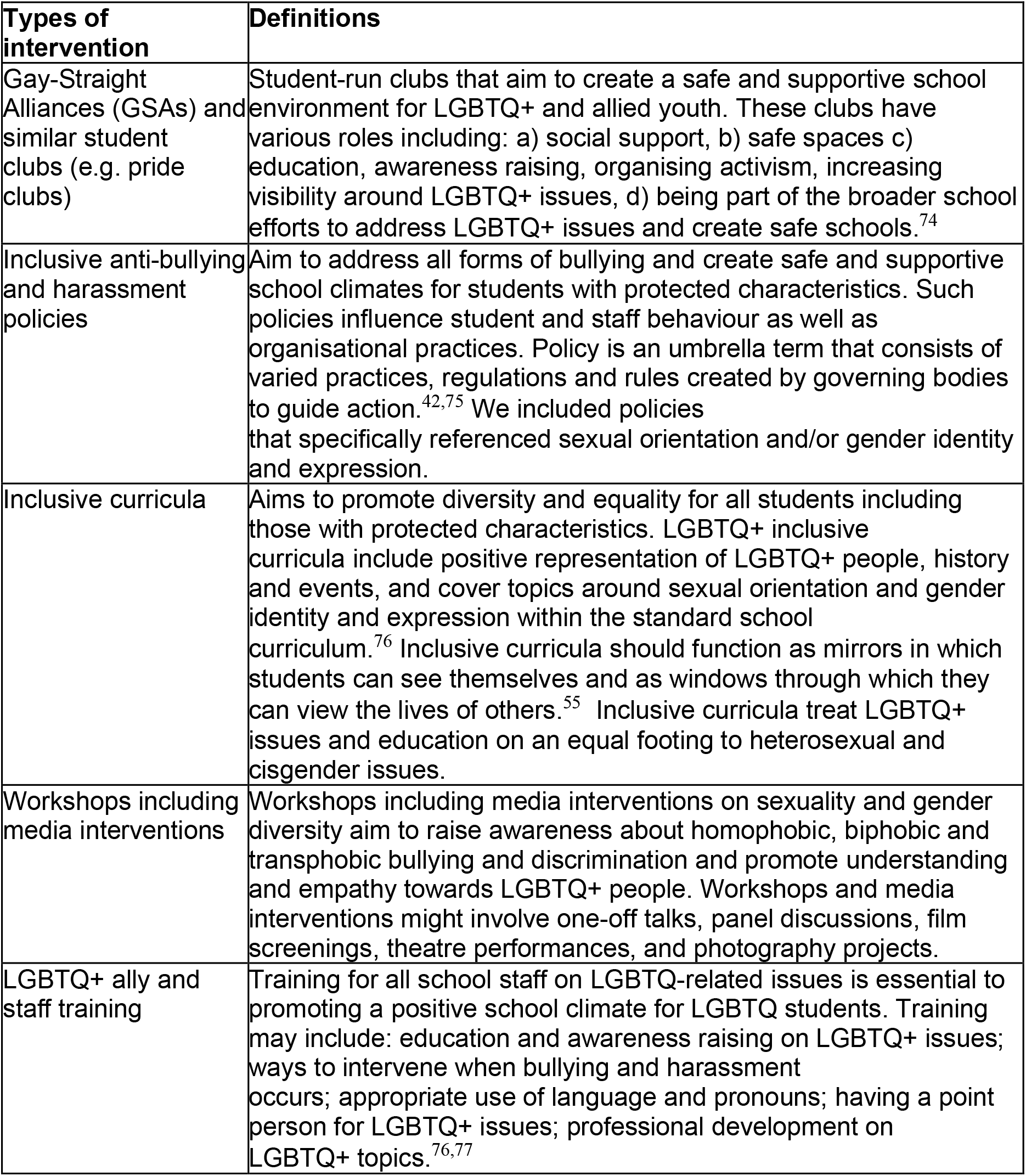
Types of interventions and what they involve.

1. Gay-Straight Alliances (GSAs) or similar student clubs (e.g. pride clubs)
2. LGBTQ+ inclusive anti-bullying and harassment policies
3. LGBTQ+ Inclusive curricula
4. Workshops including media interventions
5. LGBTQ+ ally and staff training

We present the main theory for each theme through an overarching CMO configuration (Table 4 and Figures 2-5). Some themes had multiple overarching CMO configurations to represent distinct outcomes or mechanisms. Each theme includes additional information around contexts, mechanisms, and potential harms. Where a CMO was raised by, or strongly supported by the YPAG or SAG, we reference ‘YPAG’ or ‘SAG.’ Individual CMOs and references for each section are provided in the Appendix.

**Table 4.** Summary of Context-Mechanism-Outcome (CMO) configurations comprising the programme theory for each intervention theme

| Type of intervention | Context (when the intervention works best) | Mechanism (why the intervention works) | Outcome | For whom |
| --- | --- | --- | --- | --- |
| Gay-Straight Alliances or similar student clubs (e.g. pride clubs) | Longer-established clubs<br>Clubs integrated in wider school strategy<br>Schools with positive climate<br>LGBTQ+ teachers attending the clubs and wearing rainbow yards | Reduced homophobia, improved relationships between students, empower SGM students, normalisation of being LGBTQ+ → improved school climate | Reduction in self-reported bullying and discrimination | Sexual minority and trans students |
|  |  | Reduced bullying and safe space for self-expression and social activities | Reduced likelihood of suicidal thoughts and attempts; reduced isolation and increased feelings of safety | Sexual minority and trans students |
| Inclusive anti-bullying and harassment policies | Longer-established policies<br>Policies being specific to LGBTQ+ issues<br>Supportive school leadership<br>Staff being aware and implementing policies<br>Education and support to bullies<br>Combination of multiple policies in least safe schools | Reduced homophobia → reduced bullying and stressors → improved school climate | Increased feelings of safety and higher self-esteem; Reduced likelihood of self-harm, suicidal thoughts and attempts | Sexual minority and trans students; differential effects for lesbian, gay and bisexual and transgender students |
| Workshops including media interventions | Workshops held by LGBTQ+ peer educators<br>Media interventions led | Increased empathy and understanding towards LGBTQ+ students; awareness of discrimination | Increased inclusivity and acceptance; Decreased homophobic and | Sexual minority and trans students |
|  | by LGBGTQ+ students<br>3. Included in a wider long-term commitment to inclusivity and acceptance by the school |  | transphobic bullying |  |
| LGBTQ+ ally and staff training | 1. Training on how to discuss homophobic language use and bullying<br>2. Sufficient training and resources<br>3. Training co-designed and co-delivered by LGBTQ+ staff and students | Staff more equipped to implement interventions, provide support and be inclusive towards LGBTQ+ students → increased acceptance, support, treatment, connection, and safe learning environments | Less victimisation; greater self-esteem, wellbeing and mental health | Sexual minority and trans students |
|  |  | Increased likelihood of discussing, responding to, and intervening with homophobic language use and bullying | Increased likelihood of feeling safe and less victimised | Sexual minority and trans students |
| Inclusive curricula | 1. Positive LGBTQ+ representation / role models<br>2. Avoiding "deficit and at-risk" narratives<br>3. Education on LGBTQ+ issues<br>4. Implementation at an early age | Increased understanding of experiences of LGBTQ+ people, including bullying → acceptance and normalisation of being LGBTQ+ and improved school climate | Decreased victimisation and bullying and increased intervention with bullying | Sexual minority and trans students, especially severely victimised students |

- LGBTQ+: Lesbian, Gay, Bisexual, Trans, Queer. - →: is hypothesised to lead to

**Figure 2.**
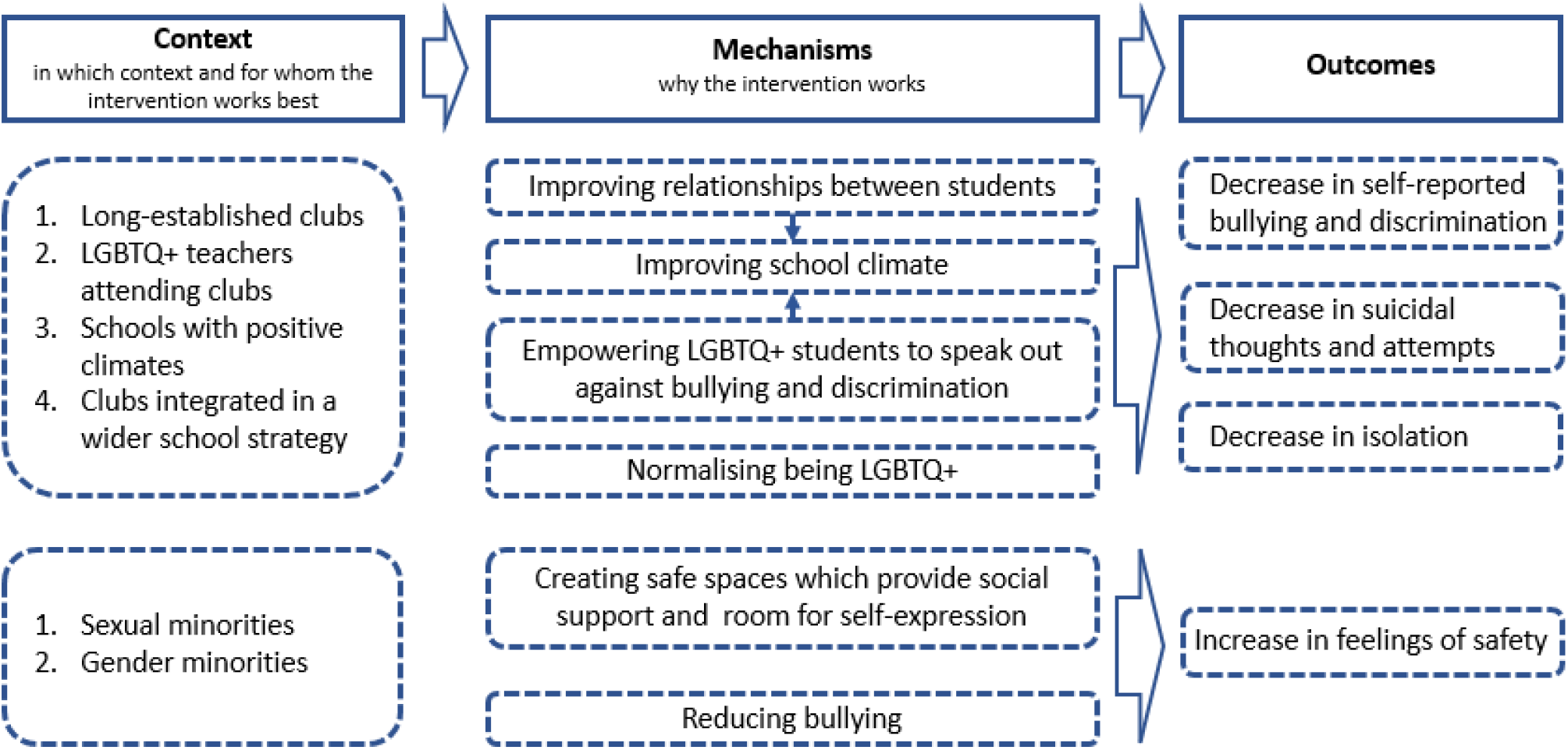
Programme theory for Gay-Straight Alliances and similar student clubs (e.g. pride clubs)

### Gay-Straight Alliances and similar student clubs (e.g. pride clubs)

#### Overarching CMO configurations (Figure 2)

When sexual and gender minority students attend schools with GSAs or similar clubs (C), they may experience reductions in bullying and discrimination (O). This could be because these clubs reduce homophobia, biphobia and transphobia in the wider school environment, improve relationships between students, empower sexual and gender minorities to speak out against bullying and discrimination, normalise being LGBTQ+, and improve the school climate (M) (SAG).^30–36^

When sexual minority and trans students attend schools with GSAs or similar clubs (C), they report reductions in suicidal thoughts and attempts, improvements in academic performance, increased school attendance, reductions in isolation, and increased feelings of safety (O). This could be because of reductions in bullying, and increases in social support and connectedness, due to creating safe spaces where students make friends, normalise their thoughts and feelings and do not feel judged, get to know/build positive relationships with school staff who are supportive of LGBTQ+ people and who they can trust (M) (SAG, YPAG).^30–33,35–38^

#### Additional information on mechanisms and strategies

When teachers who identify as sexual or gender minorities also attend GSAs and similar clubs, it may enhance their positive impact because students are exposed to role models who they can turn to for support (SAG, YPAG).^37^ Staff can communicate their support by attending GSAs or wearing rainbow lanyards (YPAG). The longer-established the GSA or similar club, the more likely it is to be effective.^33,34^ It is also important that GSAs and similar clubs are taken as seriously as other clubs (YPAG).

#### Key contexts and groups

Young people who are still coming to terms with their sexual orientation or gender may not attend GSAs or similar clubs. However, the presence of a GSA or similar club could be more important than participating in it, perhaps because the activities benefit the whole school.^30^ Setting up a successful GSA might depend on school climate including openness amongst students and staff, a whole-school “inclusivity” approach as well as tailoring the groups to the school’s demographics and ethos (SAG). Resistance and ignorance from parents, conservatism in families, lack of confidence or skills deficit in teachers as well as single-sex boys’ schools can be barriers to successfully implementing GSAs (SAG). One study found that although GSAs reduced bullying and improved feelings of safety, there was no reduction in depressive symptoms.^30^ Reasons for this finding were unclear.

#### Potential for harm

If the wider school environment is not supportive, GSAs or similar clubs could increase bullying because the visibility of sexual minority and trans students is heightened.^37,39^ Sexual minority and trans students might be reluctant to attend GSAs or similar clubs if they fear being stigmatised and bullied by other students and staff for attending them (SAG, YPAG). This might particularly be the case in rural settings.^39^ Members of GSAs, or similar clubs, might also become isolated from the wider school community (YPAG). The wider school context has to be addressed beyond club meetings and the climate of a school could be assessed first to determine what type of intervention might be most effective (YPAG, SAG). Our SAG also suggested that beyond the wider environment, if a GSA is not run well then it might not be a safe space for all members and as a result not inclusive in itself (SAG).

### Inclusive anti-bullying and harassment policies

#### Overarching CMO configurations (Figure 3)

When sexual minority and trans students attend schools with inclusive anti-bullying and harassment policies, and staff are aware of these policies and implement them (C), sexual minority and trans students feel safer, have higher self-esteem and are less likely to experience self-harm, suicidal ideation, suicide attempts and absenteeism (O). This could be because of reduced bullying and homophobic aggression^40,41^ and a more supportive school culture, with staff and students likely to intervene (M).^32,33,36,40–45^

**Figure 3.**
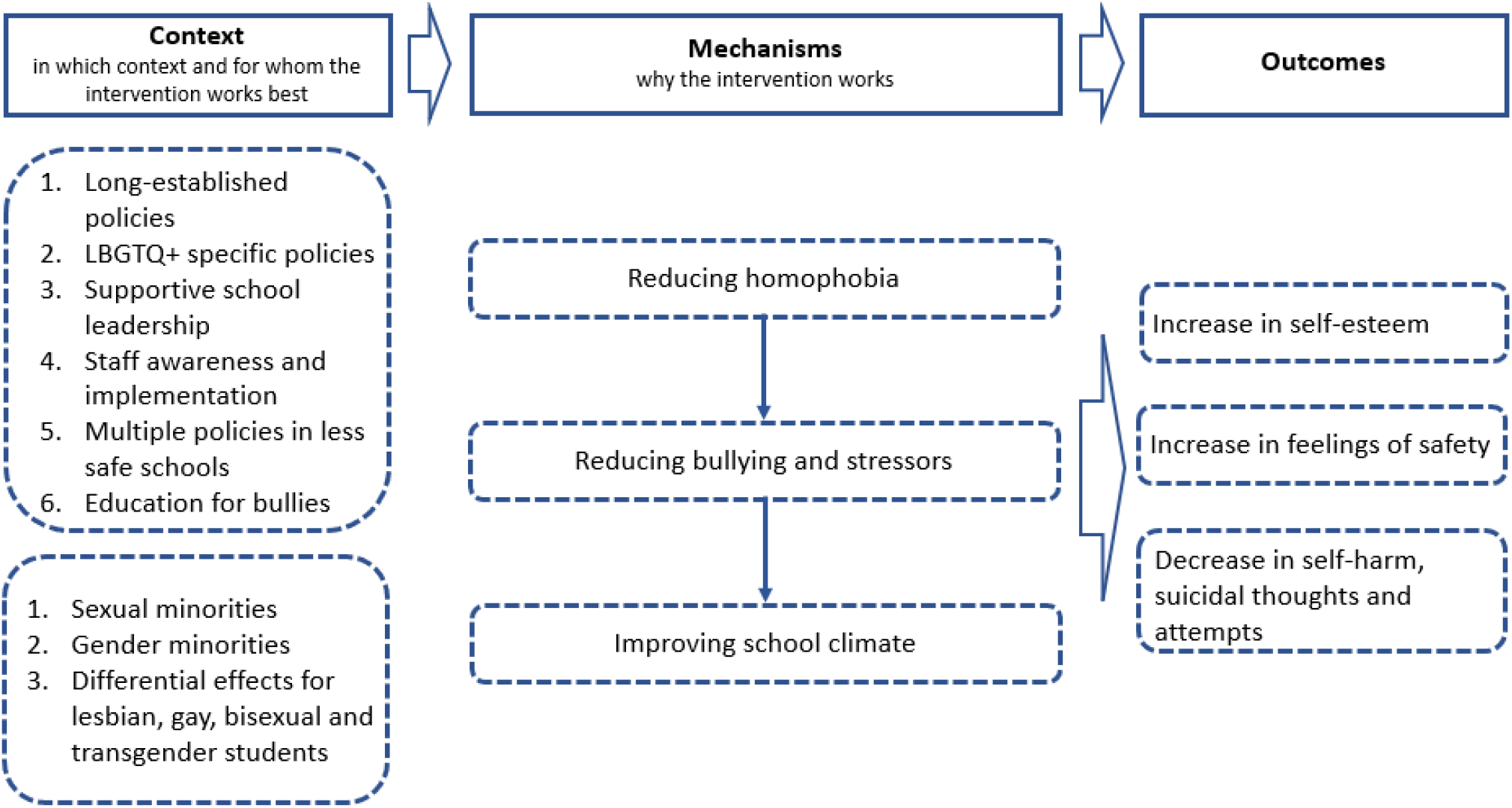
Programme theory for inclusive anti-bullying and harassment policies

When teachers and school staff implement inclusive policies in rural or politically conservative communities, with religious groups that oppose equal rights (C), they may face barriers such as unsupportive school leadership, patriarchal values, and hetero- and cisnormativity (O) due to a lack of systemic changes to attitudes (M).^46–48^

When inclusive anti-bullying policies address homophobic language within broader conversations about social status, popularity and masculinity (C), this is more likely to reduce homophobic language and slurs (O). This could be because heterosexual students often do not see themselves as homophobic, but they understand ideas about popularity and masculinity (M).^49^

#### Additional information on mechanisms and strategies

It is important that policies are supported by school leaders and the implementation of policies is monitored. If schools have processes in place to record incidents of homophobic, biphobic, and transphobic bullying, then students and teachers might be more likely to report bullying (SAG).

#### Key contexts and groups

It is possible that lesbian and gay, but not bisexual or trans, students are at reduced risk of bullying and suicide attempts in schools with inclusive anti-bullying policies compared to those without.^42^ This might be because the risk factors are different among bisexual and trans, compared with gay and lesbian young people.^42,41^ The positive effects of inclusive school policies might be less likely to persist among all boys/young men than girls/young women.^50^ It seems necessary that the school policy is an LGBTQ+ inclusive one, not just a general one, as these do not reduce bullying among sexual minority and trans students.^42^

#### Potential for harms

Gender equity government education legislation addresses gender inequity in schools. When gender equity policies are implemented in schools that are hostile to sexual and gender minorities, these students might experience increases in bullying or isolation.^47^ Students might gain a false sense of safety due to policies, and face backlash when being “out” about their sexual or gender.^51^ Our YPAG proposed conflict resolution talks to address bullying instead of punishments such as detention, which do not educate the perpetrators. They also suggested that safeguarding issues should be evaluated to respect the privacy of sexual minority and trans students (YPAG) when reporting bullying incidents. Information about students’ sexual or gender identity should not be revealed to parents/carers.^51^

### Inclusive curricula

#### Overarching CMOs (Figure 4)

When schools have inclusive curricula, with positive representation of sexual and gender minorities (C), sexual minority and trans students are less likely to be bullied, and other students are more likely to intervene (O1). This can improve connectedness among all young people (O2) as well as improve self-esteem and wellbeing and reduce suicidal ideation among sexual minority and trans young people (O3). This could be because inclusive curricula increase awareness, understanding and acceptance (M1), normalise and validate sexual and gender minorities (M2), oppose compulsory heterosexuality (M3), and improve the school climate (M4).^29,31,37,52–64^

**Figure 4.**
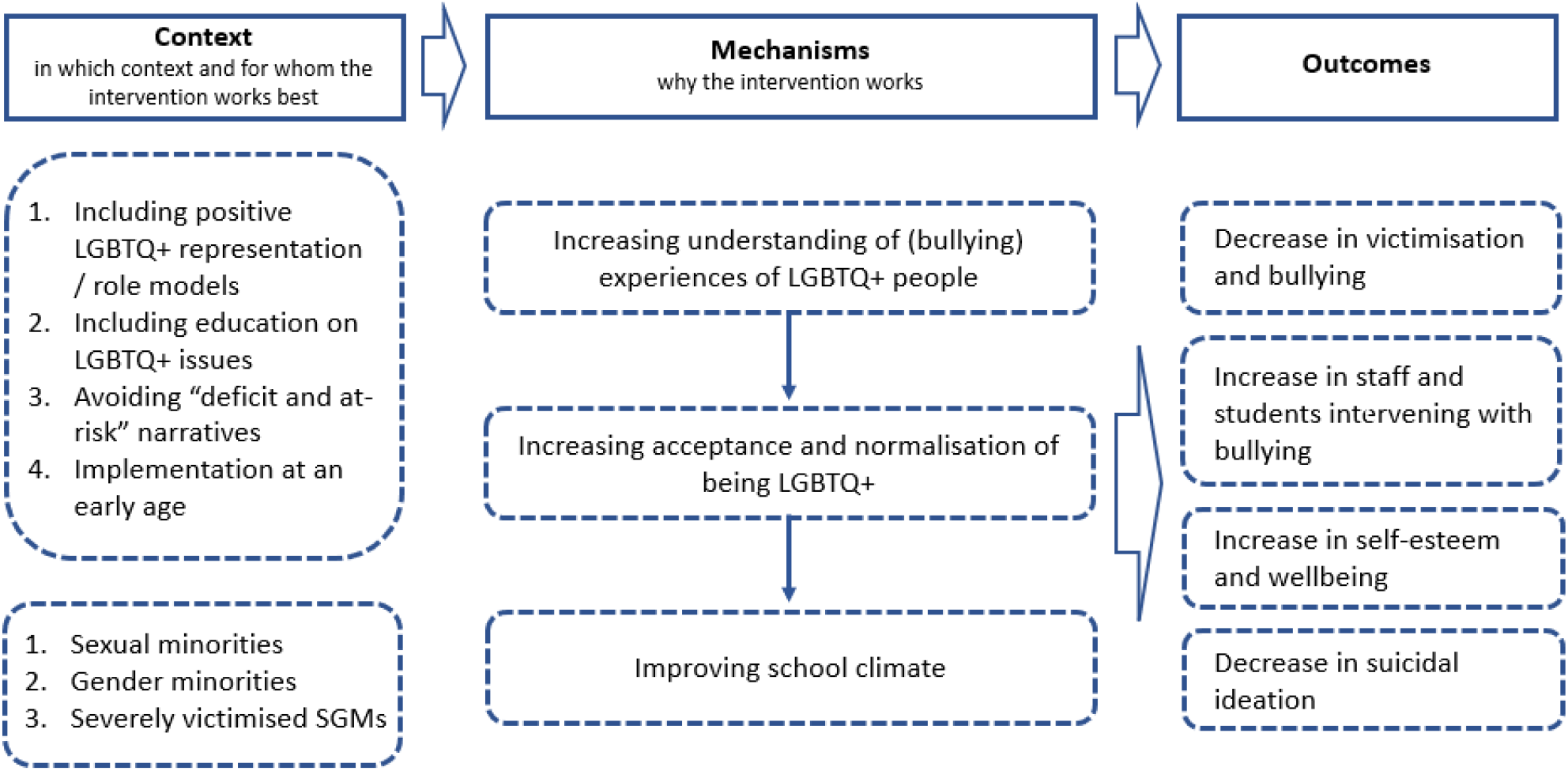
Programme theory for inclusive curricula

#### Additional information on mechanisms and strategies

Inclusive curricula seem to be most effective when they: avoid “deficit and at-risk narratives”, make the contributions and achievements of LGBTQ+ role models visible, use workbooks and literature that include LGBTQ+ issues, facilitate in-depth reflection on LGBTQ+ topics beyond learning facts, have sticker systems to highlight books with LGBT themes and/or characters, include LGBTQ+ topics in sexual health education, and are implemented from an early age onwards (SAG, YPAG).^37,52–59^ Ideally, inclusive curricula should be co-designed and co-delivered by teachers and LGBTQ+ students (SAG). Our YPAG stated that students should be better educated on the history of LGBTQ+ people, for example the lesbian community providing activism and support during the HIV/AIDS crisis. Our SAG suggested that external speakers such as mental health professional and human rights activists can provide additional insights into the challenges LGBTQ+ people experience on an everyday basis.

#### Key contexts and groups

Inclusive curricula seem to be particularly effective for students who are severely victimised based on their gender expression, or in schools with hostile climates.^31,58^ Not all studies found reductions in bullying and victimisation after implementing inclusive curricula.^29,57^ While it is unclear what the underlying mechanisms of these differential effects are, it might be due to differing school climates and specific ingredients of curricula.

#### Potential for harms

When inclusive curricula face a backlash from the wider community, they might lead to increased bullying of sexual minority and trans students.^48^ Our SAG suggested that schools might face pushback from parents who are opposed to inclusive curricula. If teachers are not well-informed on LGBTQ+ issues they might not address topics sensitively and use incorrect language and/or pronouns (SAG, YPAG). They might also fear to unintentionally cause offence (SAG).

### Workshops including media interventions

#### Overarching CMO configurations (Figure 5)

When students attend workshops on sexual and gender diversity, led by sexual and gender minorities, or assemblies and media interventions led by LGBTQ+ students (C), this increases inclusivity and acceptance towards sexual minority and trans students, decreases bullying, and increases the likelihood of students intervening against bullying (O). This could be because workshops increase students’ understanding and acceptance of sexual and gender minorities, promote empathy, and raise awareness of the harmful effects of discrimination (M) (YPAG).^60–65^

**Figure 5.**
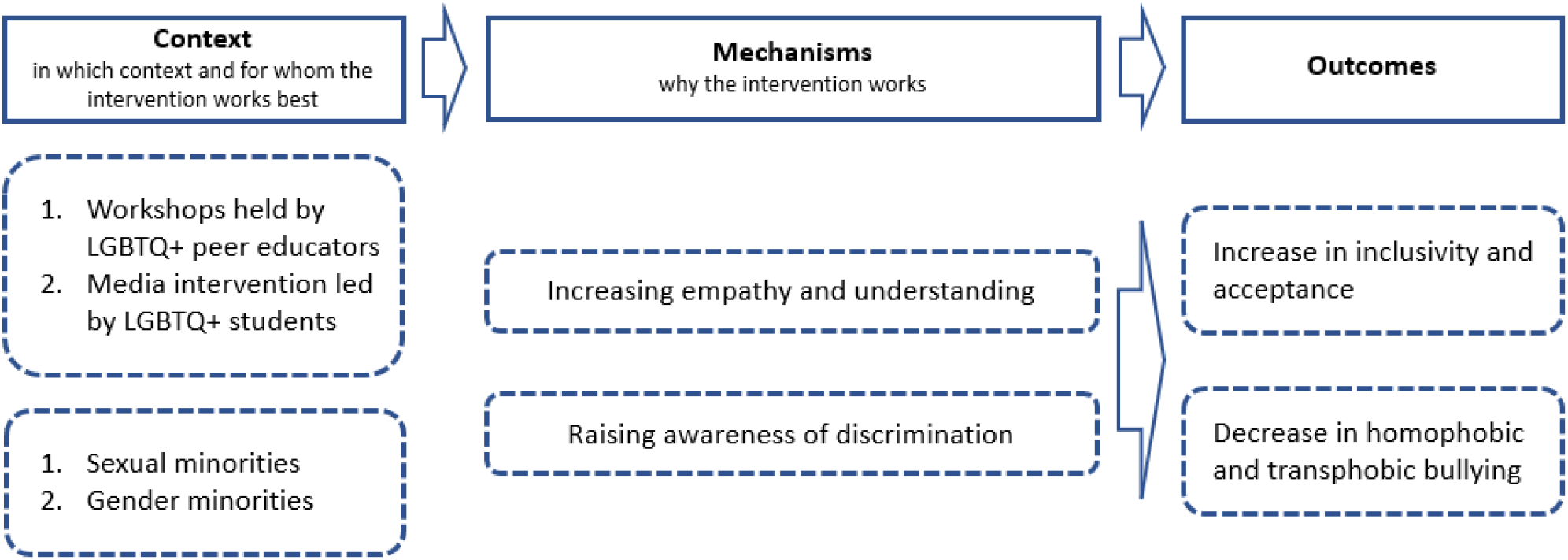
Programme theory for workshops including media interventions

#### Additional information on mechanisms and strategies

Peer educators with lived experience seem to play an important role in increasing inclusivity and acceptance and reducing bullying.^60,62,63^ Interventions might be particularly effective if they provide clear information on how to be an ally and how to behave when witnessing harassment.^64^ Young students might especially benefit from workshops and media interventions as this can foster acceptance and inclusion from a young age (SAG). However, one study in the Netherlands found only marginal and mixed effects of a peer intervention on attitudes and bullying among male students. This might be due to the content of the intervention, the school context in which the intervention was implemented, and/or the young age of students.^66^ The underlying mechanisms for this were unclear. Workshops should not be tokenistic (e.g. occurring during pride month but not thereafter) and should be part of a wider, meaningful, long-term commitment by the school including different school interventions (SAG, YPAG).

#### Potential for harms

In a study conducted in the Netherlands, there was some evidence that positive attitudes towards sexual minority and trans students, and willingness to intervene, declined after a peer-led intervention, particularly among male students. This could have been due to the content and nature of the intervention as well as the wider school context.^66^

### LGBTQ+ ally and staff training

#### Overarching CMO configurations (Figure 6)

When teachers and school staff are well-informed about sexuality and gender issues (C) sexual minority and trans students experience less victimisation, greater self-esteem, improved mental health, fewer days of school absence, and higher attainment (O). This could be because staff are better equipped to create safe spaces, support GSAs and inclusive curricula, and refer students to community and counselling support (M1). Students are also likely to build connections and feel accepted within a safe and progressive environment where gender binary norms are challenged, and staff use correct pronouns (M2).^31,54,67–69^

**Figure 6.**
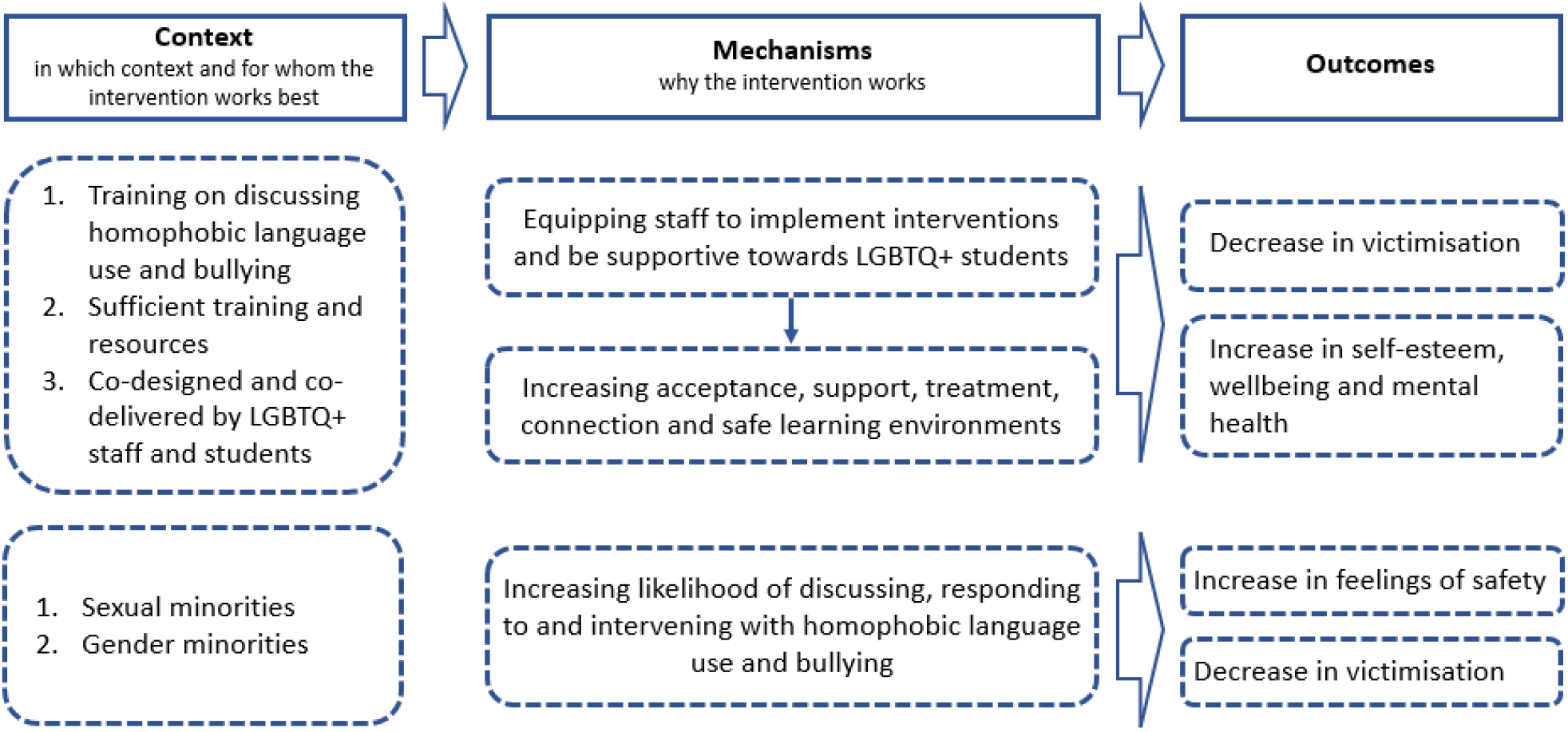
Programme theory for LGBTQ+ ally and staff training

When teachers receive training in how to be an ally, which provides them with information about language use and behaviour (C), sexual and gender minority students feel safer and less victimised (O). This could be because teachers and students are more likely to discuss, respond to and intervene against such behaviour (M).^39,67–72^

#### Additional information on mechanisms and strategies

One of the barriers to teaching staff supporting sexual minority and trans students is insufficient training and resources, including lack of knowledge about pronouns (SAG, YPAG).^68^ Teachers might be more likely to discuss homophobic language in class, but not more likely to intervene after a training course, if the course does not sufficiently prepare them to do so.^70^ Training on LGBTQ+ topics might be particularly effective if co-designed and co-delivered by teachers and LGBTQ+ students (SAG, YPAG).

## Discussion

We identified five types of universal intervention designed to promote inclusivity and acceptance of diverse sexual and gender identities in secondary schools. These interventions included Gay-Straight Alliances or similar student clubs (e.g. pride clubs), LGBTQ+ inclusive anti-bullying and harassment policies, LGBTQ+ inclusive curricula, workshops including media-based interventions, and LGBTQ+ ally and staff training. We produced a conceptual framework to explain how these interventions might work, for whom, in which contexts, and why. Consistent with minority stress theory and its supporting evidence, our hypothesis was that improving inclusivity and acceptance for sexual minority and trans young people in schools would reduce their risk of subsequent depression, anxiety, self-harm and suicidality.

### Strengths and limitations

Our literature search was systematic but, consistent with recommendations for Rapid Realist Reviews, we did not aim to capture all studies exhaustively.^27^ Also consistent with most Rapid Realist Reviews, we did not assess the methodological quality of individual studies. Instead, we assessed the relevance of each study to our programme theories, which were the main outputs of our investigation. Our programme theories were informed, refined and endorsed by experts by lived experience, including young people, teachers, policy representatives and school governors. This should improve the validity and generalisability of our theories, and the relevance and feasibility of our recommendations for policy and practice.

Although our hypotheses were generally supported, few studies reported data on depression and anxiety. Several studies reported data on self-harm and suicidality. Interventions that reduce the risk of self-harm and suicidality are also likely to affect depression and anxiety. However, more research on depression and anxiety would be beneficial.

Most studies were conducted in North America or Australia. Findings from these countries may not generalise to other settings, particularly low- and middle-income countries. Few studies were large enough to meaningfully distinguish between sexual minority or trans sub-groups. We also found little evidence on whether the effectiveness of interventions varied according to age, ethnicity, or symptom severity. We found only one Randomised Controlled Trial.^57^ Future research could investigate the effectiveness of these interventions in schools using Randomised Controlled Trials. Studies could also explore differences for sexual minority and trans students based on ethnicity, religion, disability, and other characteristics.

### Summary of findings and recommendations for schools and policy makers

Gay-Straight Alliances or similar student clubs seem to perform better when they are longer-established and attended by teaching staff who are sexual minority or trans role models. The potential benefits of Gay-Straight Alliances or similar student clubs might depend upon the pre-existing school climate. These clubs are likely to make sexual minority and trans students more visible, which could increase their exposure to bullying and discrimination. It is therefore possible that Gay-Straight Alliances and similar clubs tend to be implemented, and continued longer-term, in schools with more positive climates.

The school climate emerged as particularly important in our review. School climate is shaped by norms, beliefs, relationships (within the school and with the community), teaching and learning practices, and the organizational and physical features of the school.^72^ Inclusive curricula and anti-bullying and harassment policies might be more effective at changing the school climate than GSAs or similar student clubs. However, these three approaches to intervention seem complementary. Implementing multiple universal approaches could maximise the possibility of changing the school climate and improving outcomes for students. The order in which interventions are implemented could also be considered. Inclusive curricula and anti-bullying and harassment policies could be implemented before GSAs or similar clubs. This would demonstrate that the school promotes inclusive and accepting attitudes towards sexual and gender minorities and does not tolerate bullying based on these characteristics. The clubs would therefore be supported by a wider movement within the school, at policy level, with the support of school leadership.

Inclusive anti-bullying and harassment policies may be less effective for bisexual and trans than for lesbian or gay students. These policies may need to be adapted so they are effective for these young people. The existence of inclusive anti-bullying and harassment policies may not be sufficient to reduce discrimination and harassment towards sexual minority and trans students. Implementation seems to depend upon the awareness of teaching staff, and the active support of school leaders and the wider community. Inclusive anti-bullying and harassment policies could work best when there is education and support for bullies (e.g. restorative justice) and a combination of multiple policies, particularly in the least safe schools.

Inclusive curricula seem to work best when there is implementation at an early age and positive representation of the achievements and contributions of sexual minority and trans role models. Inclusive curricula could avoid focusing on “deficit and at-risk” narratives and normalise sexual and gender minorities as equal to heterosexual and cisgender people. The implementation and effectiveness of all interventions is likely to depend on how competent and well-trained teaching staff are with these issues. Sufficient teacher training and resources could be provided so that teachers are educated to be aware of, and feel confident at challenging, slurs and bullying. Teaching staff might then be better equipped to implement interventions, provide support, and be inclusive towards sexual minority and trans students. This could lead to increased acceptance, support, and safer learning environments. In turn this could reduce bullying and improve mental health for sexual minority and trans students. Inclusive curricula could benefit all sexual minority and trans students, especially those who have experienced severe victimisation.

Representation of sexual minority and trans role models emerged as an important theme in our review. For example, workshops and media interventions might be more effective when they are led by people who are sexual or gender minorities. This could increase empathy, awareness and understanding, and lead to increased inclusivity and acceptance.

It might be harder to reduce homophobia, biphobia and transphobia among boys and young men compared with girls and young women. This is perhaps consistent with evidence that women are less likely to hold negative attitudes towards sexual minorities than men.^73^ Universal interventions in schools could take this into account. One way that interventions could be adapted for boys is to focus less on the terms homophobia, biphobia and transphobia and instead challenge issues of masculinity and popularity.

Our findings provide guiding principles for schools to develop and implement universal interventions, which could improve inclusivity and acceptance for sexual minority and trans students, and reduce their risk of depression, anxiety, self-harm and suicidality. In line with the realist approach, our findings should also encourage primary research to confirm, refute, and refine our theories.^25^

## Data Availability

All data produced in the present study are available upon reasonable request to the authors

## Acknowledgements and funding

This work was funded by a Wellcome Trust Mental Health Active Ingredients commission awarded to GL at University College London. We would like to thank the McPin Foundation, a leading charity placing lived experience at the heart of mental health research, for recruiting our Young Person’s Advisory Group (YPAG), convening our involvement meetings, and leading our co-production work. We would also like to thank members of our stakeholder advisory group, for advising on the development of our programme theory.

## References

1 McManus S. Mental health of children and young people in England, 2017. NHS DIgital 2018.

2 Solmi M, Radua J, Olivola M, et al. Age at onset of mental disorders worldwide: large-scale meta-analysis of 192 epidemiological studies. Molecular Psychiatry 2021 2021; 17: 1–15.

3 Mars B, Heron J, Crane C, et al. Differences in risk factors for self-harm with and without suicidal intent: findings from the ALSPAC cohort. J Affect Disord 2014; 168: 407–14.

4 Mars B, Heron J, Crane C, et al. Clinical and social outcomes of adolescent self harm: population based birth cohort study. BMJ 2014; 349: g5954.

5 Morgan C, Webb RT, Carr MJ, et al. Incidence, clinical management, and mortality risk following self harm among children and adolescents: Cohort study in primary care. BMJ (Online) 2017; 359. DOI:10.1136/bmj.j4351.

6 Bould H, Mars B, Moran P, Biddle L, Gunnell D. Rising suicide rates among adolescents. Lancet 2019; 0. DOI:10.1016/S0140-6736(19)31102-X.

7 Jones A, Robinson E, Oginni O, Rahman Q, Rimes KA. Anxiety disorders, gender nonconformity, bullying and self-esteem in sexual minority adolescents: prospective birth cohort study. Journal of Child Psychology and Psychiatry 2017; 58: 1201–9.

8 Lucassen MF, Stasiak K, Samra R, Frampton CM, Merry SN. Sexual minority youth and depressive symptoms or depressive disorder: A systematic review and meta-analysis of population-based studies. Australian & New Zealand Journal of Psychiatry 2017; 51: 774–87.

9 Batejan KL, Jarvi SM, Swenson LP. Sexual orientation and non-suicidal self-injury: a meta-analytic review. Arch Suicide Res 2015; 19: 131–50.

10 Reisner SL, Poteat T, Keatley JA, et al. Global health burden and needs of transgender populations: a review. Lancet 2016; 388: 412–36.

11 Valentine SE, Shipherd JC. A systematic review of social stress and mental health among transgender and gender non-conforming people in the United States. Clinical Psychology Review. 2018; 66: 24–38.

12 Reisner SL, Biello KB, Hughto JMW, et al. Psychiatric Diagnoses and Comorbidities in a Diverse, Multicity Cohort of Young Transgender Women: Baseline Findings From Project LifeSkills. JAMA Pediatrics 2016; 170: 481–6.

13 SR V, CB B, DV G, J S. Mental Health and Psychosocial Risk and Protective Factors Among Black and Latinx Transgender Youth Compared With Peers. JAMA Netw Open 2021; 4. DOI:10.1001/JAMANETWORKOPEN.2021.3256.

14 Irish M, Solmi F, Mars B, et al. Depression and self-harm from adolescence to young adulthood in sexual minorities compared with heterosexuals in the UK: a population-based cohort study. Lancet Child Adolesc Health 2018; 3: 91–8.

15 Amos R, Manalastas EJ, White R, Bos H, Patalay P. Mental health, social adversity, and health-related outcomes in sexual minority adolescents: a contemporary national cohort study. The Lancet Child & Adolescent Health 2019; 0: 1–10.

16 Spizzirri G, Eufrásio R, Lima MCP, et al. Proportion of people identified as transgender and non-binary gender in Brazil. Scientific Reports 2021 11:1 2021; 11: 1–7.

17 Rose G. Sick Individuals and Sick Populations. 1985.

18 Doyle YG, Furey A, Flowers J. Sick individuals and sick populations: 20 Years later. Journal of Epidemiology and Community Health 2006; 60: 396–8.

19 Schuster MA, Bogart LM, Klein DJ, et al. A Longitudinal Study of Bullying of Sexual-Minority Youth. New England Journal of Medicine 2015; 372: 1872–4.

20 McDermott E, Hughes E, Rawlings V. The social determinants of lesbian, gay, bisexual and transgender youth suicidality in England: a mixed methods study. Journal of Public Health 2018; 40: e244–51.

21 Joseph Kosciw by G, Greytak EA, Giga NM, Christian Villenas M, Danischewski DJ. The 2015 National School Climate Survey. 2016.

22 McDermott, Elizabeth; Hughes, Elizabeth; Rawlings V. Queer Futures: Understanding lesbian, gay, bisexual and trans (LGBT) adolescents’ suicide, self-harm and help-seeking behaviour. 2016.

23 Meyer IH. Prejudice, social stress, and mental health in lesbian, gay, and bisexual populations: Conceptual issues and research evidence. Psychological Bulletin 2003; 129: 674–97.

24 KKH T, GJ T, SJ E, JM S, JF V. Gender Minority Stress: A Critical Review. J Homosex 2020; 67: 1471–89.

25 E D, M J. Rapid Realist Review of School-Based Physical Activity Interventions in 7-to 11-Year-Old Children. Children (Basel) 2021; 8. DOI:10.3390/CHILDREN8010052.

26 Pawson R, Tilley Nick. Realistic evaluation. 1997; : 235.

27 Saul JE, Willis CD, Bitz J, Best A. A time-responsive tool for informing policy making: Rapid realist review. Implementation Science 2013; 8: 103.

28 CD W, JE S, J B, K P, A B, B J. Improving organizational capacity to address health literacy in public health: a rapid realist review. Public Health 2014; 128: 515–24.

29 Saewyc EM. School-based interventions to reduce health disparities among LGBTQ youth: Considering the evidence. 2016.

30 Ioverno S, Belser AB, Baiocco R, Grossman AH, Russell ST. The Protective Role of Gay-Straight Alliances for Lesbian, Gay, Bisexual, and Questioning Students: A Prospective Analysis. DOI:10.1037/sgd0000193.

31 Kosciw JG, Palmer NA, Kull RM, Greytak EA. The Effect of Negative School Climate on Academic Outcomes for LGBT Youth and the Role of In-School Supports. https://doi.org/101080/153882202012732546 2012; 12: 45–63.

32 Day JK, Fish JN, Grossman AH, Russell ST. Gay-Straight Alliances, Inclusive Policy, and School Climate: LGBTQ Youths’ Experiences of Social Support and Bullying. J Res Adolesc 2020; 30 Suppl 2: 418–30.

33 Saewyc EM, Konishi C, Rose HA, Homma Y. School-Based Strategies to Reduce Suicidal Ideation, Suicide Attempts, and Discrimination among Sexual Minority and Heterosexual Adolescents in Western Canada. Int J Child Youth Family Stud 2014; 5: 89.

34 Konishi C, Saewyc E, Homma Y, Poon C. Population-level evaluation of school-based interventions to prevent problem substance use among gay, lesbian and bisexual adolescents in Canada. Prev Med (Baltim) 2013; 57: 929–33.

35 Mayberry M, Chenneville T, Currie S. Challenging the Sounds of Silence: A Qualitative Study of Gay–Straight Alliances and School Reform Efforts. http://dx.doi.org/101177/0013124511409400 2011; 45: 307–39.

36 Black WW, Fedewa AL, Gonzalez KA. Effects of “Safe School” Programs and Policies on the Social Climate for Sexual-Minority Youth: A Review of the Literature. http://dx.doi.org/101080/193616532012714343 2012; 9: 321–39.

37 Harris R, Wilson-Daily AE, Fuller G. ‘I just want to feel like I’m part of everyone else’: how schools unintentionally contribute to the isolation of students who identify as LGBT+. https://doi.org/101080/0305764X20211965091 2021. DOI:10.1080/0305764X.2021.1965091.

38 Johns MM, Poteat VP, Horn SS, Kosciw J. Strengthening Our Schools to Promote Resilience and Health Among LGBTQ Youth: Emerging Evidence and Research Priorities from The State of LGBTQ Youth Health and Wellbeing Symposium. LGBT Health 2019; 6: 146–55.

39 De Pedro KT, Lynch RJ, Esqueda MC. Understanding safety, victimization and school climate among rural lesbian, gay, bisexual, transgender, and questioning (LGBTQ) youth. https://doi.org/101080/1936165320181472050 2018; 15: 265–79.

40 Jones TM, Hillier L. Sexuality education school policy for Australian GLBTIQ students. Sex Education 2012; 12: 437–54.

41 Day JK, Ioverno S, Russell ST. Safe and supportive schools for LGBT youth: Addressing educational inequities through inclusive policies and practices. Journal of School Psychology 2019; 74: 29–43.

42 Hatzenbuehler ML, Keyes KM. Inclusive Anti-bullying Policies and Reduced Risk of Suicide Attempts in Lesbian and Gay Youth. Journal of Adolescent Health 2013; 53: S21–6.

43 Day JK, Snapp SD, Russell ST. Supportive, Not Punitive, Practices Reduce Homophobic Bullying and Improve School Connectedness. 2016. DOI:10.1037/sgd0000195.supp.

44 Russell ST, Day JK, Ioverno S, Toomey RB. Are school policies focused on sexual orientation and gender identity associated with less bullying? Teachers’ perspectives. Journal of School Psychology 2016; 54: 29–38.

45 Green AE, Willging CE, Ramos MM, Shattuck D, Gunderson L. Factors Impacting Implementation of Evidence-Based Strategies to Create Safe and Supportive Schools for Sexual and Gender Minority Students. J Adolesc Health 2018; 63: 643–8.

46 Steck AK, Perry D. Challenging Heteronormativity: Creating a Safe and Inclusive Environment for LGBTQ Students. https://doi.org/101080/1538822020171308255 2017; 17: 227–43.

47 Sinacore AL, Chao SC, Ho J. Gender Equity Education Act in Taiwan: influences on the school community. International Journal for Educational and Vocational Guidance 2018 19:2 2018; 19: 293–312.

48 Ginicola M, Smith C, Rhoades E. Love Thy Neighbor: A Guide for Implementing Safe School Initiatives for LGBTQ Students in Nonaffirming Religious Communities. Journal of LGBT Issues in Counseling 2016; 10: 159–73.

49 Fulcher K. That’s so homophobic? Australian young people’s perspectives on homophobic language use in secondary schools. https://doi.org/101080/1468181120161275541 2017; 17: 290–301.

50 Van de Ven P. Effects on High School Students of a Teaching Module for Reducing Homophobia. http://dx.doi.org/101080/0197353319959646137 2011; 17: 153–72.

51 Moyano N, Sánchez-Fuentes M del M. Homophobic bullying at schools: A systematic review of research, prevalence, school-related predictors and consequences. Aggression and Violent Behavior 2020; 53: 101441.

52 Francis DA. What does the teaching and learning of sexuality education in South African schools reveal about counter-normative sexualities? https://doi.org/101080/1468181120181563535 2018; 19: 406–21.

53 Karla Fleshman R. Building Resilience, Reducing Risk: Four Pillars to Creating Safer, More Supportive Schools for LGBTQ+ Youth. DOI:10.32481/djph.2019.06.009.

54 Evans I, Rawlings V. “It was Just One Less Thing that I Had to Worry about”: Positive Experiences of Schooling for Gender Diverse and Transgender Students. https://doi.org/101080/0091836920191698918 2019; 68: 1489–508.

55 Baams L, Dubas JS, van Aken MAG. Comprehensive Sexuality Education as a Longitudinal Predictor of LGBTQ Name-Calling and Perceived Willingness to Intervene in School. Journal of Youth and Adolescence 2017; 46: 931–42.

56 Rabbitte M. Sex Education in School, are Gender and Sexual Minority Youth Included?: A Decade in Review. Am J Sex Educ 2020; 15: 530.

57 Espelage DL, Low S, van Ryzin MJ, Polanin JR. Clinical Trial of Second Step Middle School Program: Impact on Bullying, Cyberbullying, Homophobic Teasing, and Sexual Harassment Perpetration. https://doi.org/1017105/spr-15-00521 2019; 44: p464–79.

58 O’Farrell M, Corcoran P, Davoren MP. Examining LGBTI+ inclusive sexual health education from the perspective of both youth and facilitators: a systematic review. BMJ Open 2021; 11: e047856.

59 Vilkin E, Einhorn L, Satyanarayana S, Eisu A, Kimport K, Flentje A. Elementary Students’ Gender Beliefs and Attitudes Following a 12-Week Arts Curriculum Focused on Gender. https://doi.org/101080/1936165320191613282 2019; 17: 70–88.

60 Burford J, Lucassen MFG, Hamilton T. Evaluating a gender diversity workshop to promote positive learning environments. https://doi.org/101080/1936165320161264910 2017; 14: 211–27.

61 Douglas N, Kemp S, Aggleton P, Warwick I. The Role of External Professionals in Education about Sexual Orientation-towards good practice. Sex Education 2001; 1: 149–62.

62 Eick U, Rubinstein T, Hertz S, Slater A. Changing attitudes of high school students in Israel toward homosexuality. https://doi.org/101080/1936165320151087930 2016; 13: 192–206.

63 Lucassen MFG, Burford J. Educating for diversity: an evaluation of a sexuality diversity workshop to address secondary school bullying. undefined 2015; 23: 544–9.

64 Wernick LJ, Dessel AB, Kulick A, Graham LF. LGBTQQ youth creating change: Developing allies against bullying through performance and dialogue. Children and Youth Services Review 2013; 35: 1576–86.

65 Wernick LJ, Kulick A, Dessel AB, Graham LF. Theater and Dialogue to Increase Youth’s Intentions to Advocate for LGBTQQ People. Research on Social Work Practice 2016; 26: 189–202.

66 Kroneman M, Admiraal W, Ketelaars M. A peer–educator intervention: Attitudes towards LGB in prevocational secondary education in the Netherlands. https://doi.org/101080/1936165320181531101 2018; 16: 62–82.

67 Mitton-Kukner J, Kearns LL, Tompkins J. Pre-service educators and anti-oppressive pedagogy: interrupting and challenging LGBTQ oppression in schools. http://dx.doi.org/101080/1359866X20151020047 2015; 44: 20–34.

68 Swanson K, Gettinger M. Teachers’ knowledge, attitudes, and supportive behaviors toward LGBT students: Relationship to Gay-Straight Alliances, antibullying policy, and teacher training. https://doi.org/101080/1936165320161185765 2016; 13: 326–51.

69 Ollis D. ‘I haven’t changed bigots but …’: reflections on the impact of teacher professional learning in sexuality education. http://dx.doi.org/101080/14681811003666523 2010; 10: 217–30.

70 Poteat VP, Slaatten H, Breivik K. Factors associated with teachers discussing and intervening against homophobic language. Teaching and Teacher Education 2019; 77: 31–42.

71 Ioverno S, Nappa MR, Russell ST, Baiocco R. Student Intervention Against Homophobic Name-Calling: The Role of Peers, Teachers, and Inclusive Curricula. J Interpers Violence 2021. DOI:10.1177/08862605211042817.

72 Maxwell S, Reynolds KJ, Lee E, Subasic E, Bromhead D. The impact of school climate and school identification on academic achievement: Multilevel modeling with student and teacher data. Frontiers in Psychology 2017; 8: 2069.

73 Swaless K, Attar TE. Moral issues; sex, gender, identiy and euthanasia. British Social Attitudes: the 34th report. 2017.

74 Griffin P, Lee C, Waugh J, Beyer C. Describing Roles that Gay-Straight Alliances Play in Schools. http://dx.doi.org/101300/J367v01n03_03 2008; 1: 7–22.

75 Hall W. The effectiveness of policy interventions for school bullying: A systematic review. J Soc Social Work Res 2017; 8: 45–69.

76 Russell ST, Bishop MD, Saba VC, James I, Ioverno S. Promoting School Safety for LGBTQ and All Students. Policy Insights Behav Brain Sci 2021; 8: 160–6.

77 Gower AL, Forster M, Gloppen K, et al. School Practices to Foster LGBT-Supportive Climate: Associations with Adolescent Bullying Involvement. Prevention Science 2017; : 1–9.

