## Supplement for "Promoting inclusivity and acceptance of diverse sexual and gender identities in schools: a Rapid Realist Review of universal interventions to improve mental health"

### APPENDIX

#### Table of contents

| **Item** | **Description** | **Page number** |
| --- | --- | --- |
| Full insights from the YPAG^a^ | Insights separated according to each intervention theme | 2 |
| Full insights from the SAG^b^ | Insights separated according to each intervention theme | 5 |
| Supplementary Table 1 | Search terms | 7 |
| Supplementary Table 2 | Characteristics of included studies | 9 |
| Supplementary Table 3 | Individual CMOs^c^ for Gay-Straight Alliances or similar student clubs (e.g. pride clubs) | 19 |
| Supplementary Table 4 | Individual CMOs^c^ for inclusive anti-bullying and harassment policies | 23 |
| Supplementary Table 5 | Individual CMOs^c^ for inclusive curricula | 30 |
| Supplementary Table 6 | Individual CMOs^c^ for One-off workshops and media interventions | 35 |
| Supplementary Table 7 | Individual CMOs^c^ for ally and staff training | 39 |

^a^Young Person’s Advisory Group

^b^Stakeholders Advisory Group

^c^Context-Mechanism-Outcome configurations

### Insights from the YPAG

The text presented here represents notes made during meetings and interactions with our YPAG, and is not written in a formal style to avoid changing the meaning.

#### Gay-Straight Alliances or similar student clubs (e.g. pride clubs)

- Clubs are only viewed as helpful when the wider school environment is positive about them. If not, sexual and gender minorities are likely to be made fun of or bullied for attending the clubs.
- Before applying interventions into schools there could be work done to assess the school (for example: how supported LGBTQ+ students feel) to then determine what interventions are administered.
- As part of pride club some of the teachers attended pride clubs and had rainbow lanyards and that was helpful as it made it clear who we could talk to about stuff. It would be good if there was a common room for sexual and gender minority students to use to recuperate or go to have a space to speak to other people. Sometimes in schools, you know which staff members you can talk to but it’s not always possible to find them. It’s important to know that there’s a place that you can get support.
- A lot of the issues with things that are implemented is that there isn’t much follow-through and the clubs aren’t taken as seriously as other clubs.
- One YP said a lot of people in their school might not be out or are still coming to terms with their sexuality so, for example, they wouldn’t go to a GSA and this should be considered.

#### Anti-bullying and harassment policies

- Punishment (e.g. detentions) for bullying should include some form of education or way of ensuring the people are learning from their mistakes. Otherwise, it can make people worse (ie escalate the bullying behaviour) which can make it worse for the victim of the situation. Bullies should talk about their feelings too and we should try to change their view.
- Chat: My sister is a primary school teacher and when students fight, they go to ‘conflict resolution’ to talk it through instead of detention.
- Safeguarding issues should be re-evaluated. Teachers need to respect the privacy of students with their parents and what information to reveal.
  - E.g. if parents get a letter home about a bullying incident and it reveals personal information about people’s sexual or gender identity, this can put students in a difficult or even dangerous situation at home.
- Being bullied about your sexual or gender identity feels different and more personal to being bullied for other reasons so we need inclusive policies.

#### Inclusive curricula

- One YP said when there are debates in a class it can end up with your peers debating things that are emotional/difficult for the queer people in the class and not treated sensitively by the cis-het students.
- We should strive towards starting discussion and learning about LGBTQ+ topics from a sensitive/positive place to cause less harm.
- One YP spoke about positive representation in their school: History teacher has a board in her classroom of people who have done great things throughout History and quite a few of them are LGBTQ+ people and the YP finds this positive.
- One YP said that what they learnt about lesbians helping during the HIV/AIDS crisis was something they learnt themselves (possibly from the TV show Pose), they didn’t learn it from school and it’s relevant History that everyone should know about
- YP said in their PSHE/PD lessons they learn about racism and discrimination but not about negative stereotypes of LGBTQ+ people, but there should be a place for this
- Teachers need to be well-informed. For example, one YP had a talk about hate crime but used incorrect words when talking LGBTQ+ people. This wasn’t done to be offensive, it was said because the teacher wasn’t educated.
- One YP said that their school has a lot of diverse books including queer authors and it makes them feel included.
- One YP said that the library in their school was a safe space for a lot of students and would really like it if their school introduced a sticker system to identify books with LGBTQ+ characters and authors.
- Another YP thought the sticker system for books is a great idea because it can help LGBTQ+ young people educate themselves and read positive stories about LGBTQ+ people.
- One YP suggested presenting positive videos to classes and schools such as sketches that include gay families to normalise this model of family life.
- Currently in classes like sex education, even if people ask about queer sex education the teachers can be hesitant to cover it, or just say that we don’t need to talk about it. This can be really difficult for teenagers as they can be quite impressionable

#### Workshops including media interventions

- One YP had a positive experience in schools but this was because of the students. The LGBTQ+ students lead assemblies to educate students, and the YP felt it was positive that this is student-led. Student-led interventions could benefit LGBTQ+ youth.
- Individual tutor groups would be a better environment to have discussions about discrimination rather than big gatherings like assemblies
- Sometimes schools can have good intentions to do LGBT specific things like events for Pride month or series of lessons. But if there isn’t effort put into it, these ideas can be dropped and ultimately give false hope for the people who wanted them to happen.
- Schools often have writing competitions. Perhaps a writing competition where people need to write a story with a queer character. This ties in with the idea of doing things that engage with the media. And it can be something that all students do. This can also apply to other minorities to improve inclusivity
- Workshops should be tokenistic or a one-off during pride month. The school should show they really care and are committed long term

#### LGBTQ+ ally and staff training

- Partnership/agreement between students and teachers to deliver training and education together about gender-related issues and LGBTQ+ topics.
- Teachers benefit from sensitivity training e.g. it is really hard for YPs when teachers use their wrong pronouns but sometimes teachers just do not know that this is important

### Insights from the SAG

The text presented here represents notes made during meetings and interactions with our SAG and is not written in a formal style to avoid changing the meaning.

#### Gay-Straight Alliances or similar student clubs (e.g. pride clubs)

- GSAs align better with secondaries and colleges but might encounter more resistance in primary and specialist sectors. Especially where cultural and religious beliefs conflict with LGBTQ+ lifestyles.
- A positive benefit of running these clubs is improved relationships between students and students with staff. CYP and staff report increased trust and respect which can positively impact CYP wellbeing and consequently their learning/progress.
- A potential mechanism in how GSAs reduce bullying and discrimination might be by promoting normalisation and increasing familiarity.
- One SAG suggested that if a GSA club is not run well then it might not be a safe space for all members and as a result not inclusive within itself.
- The successful set up of a GSA depends on school climate and tailoring the groups to the school's demographics/ethos.
- The current social justice movement might facilitate the successful set up of GSAs in that pupils might feel the topics are given sufficient national attention to be discussed without it being too personal to them individually.
- Integrating topis of equality and diversity across the school activities and a change in the school’s central value to “Inclusivity” can result in a whole-school approach which can be effective.
- Getting pupils to see the same faces each week so that they have friendly faces around school across year groups. This also allows them to come out more easily in a safer microcosm to each other and work out their identity before telling teachers, families and non-LGBT+ friends.
- Barriers: parental fear/ignorance; conservatism in families; teacher skills deficit or lack of confidence; single-sex boys' schools.
- Facilitators: strong CYP participation; effective coaching and professional learning; communications strategy to parents/community; positive role models; social justice movement.

#### Anti-bullying and harassment policies

- Individual school context applies and needs consideration. Religious schools or those still governed in a patriarchal manner will need more coaxing and education/support.
- Anti-bullying and harassment policies can’t exist in isolation. They need to be backed up by practice on the ground. It’s not just the anti-bullying and harassment policy, it’s also grievances and complaints, use of social media and home-school agreement territory.
- If a school updates its internal records system for recording incidents to have homophobic, biphobic and transphobic (HBT) categories, then it is likely in the short term that there is more HBT peer-on-peer abuse recorded as the culture is becoming more open.
- Barriers: Lack of senior leader support/governing body support; Not implementing existing policies and as such not affecting positive change; consistency of following policy and process guidance.
- Facilitators: Initial Teacher Education (ITE) and National Professional Qualification in Headship (NPQH) training covers this; Professional Teaching Standards and EWC Code of Conduct set clear expectations of teaching professionals; embedding this in overall children’s rights approach to education.

#### Inclusive curricula

- Inclusive curricula should facilitate in-depth reflection on LGBTQ+ topics and reasons for discussing such topics rather than just learning facts about influential LGBTQ+ people.
- Inclusive curricula can make students feel more connected to their peers as they are able to understand their differences and communalities. This can improve wellbeing and self-esteem.
- It can be helpful to discuss inspirational and accessible role models and use positive, reinforcing language, e.g. in form of celebrating diversity rather than discussing “struggles” or focusing on “otherness”.
- Inclusive curricula should be co-designed and co-constructed to include students’ voice and ensure that they are not tokenistic.
- It can be helpful to use the metaphor of a “mirror” (signifying that all students would see themselves in our curriculum) and a “window” (representing our ambition to show all students the world beyond their immediate experience).
- External speakers can provide additional insights into challenges experienced by LGBTQ+ people. This can include human rights activists, lawyers, etc.
- Barriers to successfully implementing inclusive curricula can include resistance and ignorance of parents, teachers, and the larger school community. Teachers who lack knowledge and confidence might fear to unintentionally cause offence.

#### One-off workshops including media interventions

- It is important that workshops are not used as a tokenistic tick box exercise.
- Workshops might be particularly impactful for younger students to sow seeds of acceptance and connectedness at an early age.
- Factors that might facilitate the impact of workshops can include increasing teacher training to build skills and confidence and incorporating the workshops in a whole school approach of embedding children’s rights. It is helpful if students can identify with and relate to role models. The long-term visibility of LGBTQ+ role models should be ensured.
- Barriers to putting on impactful workshops might include resistance and ignorance of parents, teachers, and the larger school community.

#### LGBTQ+ ally and staff training

- Staff training can increase staff’s ability and confidence in creating safe spaces by building understanding and skills.
- Barriers to successful staff training might include a lack of resources and supplies, resistance among parents, teachers, and the wider school community due to a fear of causing offence, but also the wider political climate and a lack of government backing for a specific organisation or training provider.
- The government needs to prioritise funding for adequate training throughout the system and engagement with all stakeholders. Staff training needs to be tied in with a whole school approach to mental health and wellbeing, with Children’s Rights Based education system, and into the performance training series and performance management cycle.

### Supplementary Table 1: Search Terms

| **PICO** | **Search Terms** |
| --- | --- |
| **Population** | LGBTQ+, LGBT*, LGB*, queer, sexual identit*, sexual orientation, gender identit*, lesbian, gay, bisexual, transgender, nonbinary, non-binary, asexual, pansexual, sexualit*, intersex, omnisexual, “questioning sexuality”, “questioning gender”, demisexual, aromantic |
| **Intervention** | School or school-based or educat* AND intervent* or program*or policy or curriculum |
| **Outcomes** | N/A |
| **Comparison** | N/A |

**Records identified through database searching (n = 5,155)**

A. **Pubmed (14.09.2021), 1,678 hits**

("lgbtq+"[Title/Abstract] OR "lgbt*"[Title/Abstract] OR "lgb*"[Title/Abstract] OR "homosexual*"[Title/Abstract] OR "queer"[Title/Abstract] OR "sexual identit*"[Title/Abstract] OR "sexual orientation"[Title/Abstract] OR "gender identit*"[Title/Abstract] OR "lesbian"[Title/Abstract] OR "gay"[Title/Abstract] OR "bisexual*"[Title/Abstract] OR "transgender"[Title/Abstract] OR "nonbinary"[Title/Abstract] OR "non-binary"[Title/Abstract] OR "asexual*"[Title/Abstract] OR "pansexual*"[Title/Abstract] OR "sexualit*"[Title/Abstract] OR "intersex"[Title/Abstract] OR "omnisexual*"[Title/Abstract] OR "demisexual*"[Title/Abstract] OR "aromantic"[Title/Abstract]) AND ("school intervent*"[Title/Abstract] OR "school based intervent*"[Title/Abstract] OR "school-based intervent*"[Title/Abstract] OR "education intervent*"[Title/Abstract] OR "educational intervent*"[Title/Abstract] OR "school program*"[Title/Abstract] OR "school based program*"[Title/Abstract] OR "school-based program*"[Title/Abstract] OR "education program*"[Title/Abstract] OR "educational program*"[Title/Abstract] OR "school polic*"[Title/Abstract] OR "school-based polic*"[Title/Abstract] OR "school based polic*"[Title/Abstract] OR "school curricul*"[Title/Abstract] OR "school-based curricul*"[Title/Abstract] OR "school based curricul*"[Title/Abstract] OR "curricul*"[Title/Abstract])

B. **Web of Science (14.09.2021), 1,272 hits**

<https://www.webofscience.com/wos/woscc/summary/22366e8d-7608-4a28-86cb-acaaf28d112b-0853e2a9/relevance/1>

(AB=("school intervention*" or "school based intervention*" or "school-based intervention" or "education* intervention*" or "school program*" or "school-based program*" or "school based program*" or "education* program*" or "school polic*" or "school based polic*" or "school-based polic*" or "education* polic*" or "school curricul*" or "school based curricul*" or "school-based curricul*" or "education* curricul*")) AND AB=(LGBTQ+ or LGBT* or LGB* or homosexual* or queer or "sexual identit*" or "sexual orientation" or "gender identit*" or lesbian or gay or bisexual* or transgender or nonbinary or non-binary or asexual* or pansexual* or sexualit* or intersex or omnisexual* or "questioning sexuality" or "questioning gender" or demisexual* or aromantic)

C. **PsycINFO (14.09.2021), 2,205 hits**

| 1. (LGBTQ+ or LGBT* or LGB* or homosexual* or queer or sexual identit* or sexual orientation or gender identit* or lesbian or gay or bisexual* or transgender or nonbinary or non-binary or asexual* or pansexual* or sexualit* or intersex or omnisexual* or questioning sexuality or questioning gender or demisexual* or aromantic).mp. [mp=title, abstract, heading word, table of contents, key concepts, original title, tests & measures, mesh] | 98815 |
| --- | --- |
| 2. (school intervention* or school based intervention* or school-based intervention or education* intervention* or school program* or school-based program* or school based program* or education* program* or school polic* or school based polic* or school-based polic* or education* polic* or school curricul* or school based curricul* or school-based curricul* or education* curricul*).mp. [mp=title, abstract, heading word, table of contents, key concepts, original title, tests & measures, mesh] | 102240 |
| 3. 1 and 2 | 2231 |
| 4. limit 3 to abstracts | 2205 |

### Supplementary Table 2. Characteristics of included studies

| **Author and year** | **Country** | **Study Aim** | **Study design** | **Sample (n)** | **Type of intervention** |
| --- | --- | --- | --- | --- | --- |
| Baams et al. 2017 | Netherlands | 1) To provide an overview of the content of sexuality education in six Dutch High schools 2) To examine whether the content or extensiveness of sexuality education at the beginning of the school year is related to a decrease in LGBTIQ name-calling and an increase in the willingness to intervene when witnessing LGBTQ name-calling at the end of the school year | Quantitative | Dutch adolescents (n=601) | Inclusive curricula |
| Bellini 2012 | Canada | The purposes of this study are to examine whether gay and lesbian students are receiving support in the public education system and how educators are trained to give this support | Qualitative | Individuals with experience in GSAs and working with students in a GSA (n=7) | Inclusive anti-bullying and harassment policies |
| Black et al. 2012 | USA | To review literature on GSAs and Safe School Act | Review | LGB youth | GSA and similar student clubs |
| Burford et al. 2017 | Aotearoa/New Zealand | To evaluate the potential of a 60-min gender diversity workshop to address bullying and  promote positive environments for learning. | Mixed methods | Secondary students (n=237) | One-off workshops and media interventions |
| Burk et al. 2018 | Canada | to evaluate the Out in Schools film-based intervention and its association with mental health outcomes and bullying experienced by sexual minority adolescents | Quantitative | 7-12-grade school students (n=29,832) | Inclusive curricula a |
| Day et al. 2016 | USA | To examine punitive and supportive policies and practices in relation to homophobic bullying and school connectedness | Quantitative | Students aged 10-18 (n=745) | Inclusive anti-bullying and harassment policies |
| Day et al. 2019 | USA | To investigate youth's experiences of general victimisation and bullying due to sexual orientation or gender (SOG-bullying), truancy, academic success, and perceptions of school climate in relation to presence of SOGI-focused policies. | Quantitative | Students aged 10-18 (n=113,148) | Inclusive anti-bullying and harassment policies |
| Day et al. 2020 | USA | To investigate whether GSAs and school policies, independently and mutually, are associated with less bullying and youths' perceptions of support from classmates and teachers. | Quantitative | LGBTQ youth aged 15-21 (n=1,061) | GSA and similar student clubs |
| De Pedro et al. 2018 | USA | To explore the relationships between LGBTQ affirming school climates and the safety and victimisation of LGBTQ students | Quantitative | High school students (n=611) | GSA and similar student clubs |
| Douglas et al. 2010 | England | To highlight opportunities for external practitioners to conduct education in schools about sexual orientation and identity to outline a useful approach to this work and to identify learning that can be drawn from this experience to further develop education about sexual orientation and identity with young people in schools. | Qualitative | Students (n=408) and teachers (n=4) | One-off workshops and media interventions |
| Eick et al. 2016 | Israel | To examine whether this activity, carried out in Israeli high schools, resulted in a change in participants' attitudes. | Quantitative | 9-11th-grade students (n=272) | One-off workshops and media interventions |
| Espelage et al. 2019 | USA | To examine the impact of the Second Step Middle School Program on homophobic name calling, bullying, sexual harassment | Quantitative | 11 and 12-year-old secondary school students (n=3,651) | Inclusive curricula |
| Espelage et al. 2019b | USA | To review studies that focused on protective factors associated with homophobic bullying perpetration and victimization among children and adolescents | Review | NA | GSA and similar student clubs; inclusive anti-bullying and harassment policies; inclusive curricula |
| Evans and Rawlings 2021 | Australia | To explore the positive experiences of trans and gender diverse students at school and investigate what leads to a safe and positive learning environment | Qualitative | Transgender/gender diverse young people aged 17-26 (n=3) | Inclusive anti-bullying and harassment policies; inclusive curricula; LGBTQ+ staff training |
| Fleshman 2019 | USA | To focus on four key areas where schools and school districts may implement changes to create safer more supportive schools for LGBTQ+ students. | Other (report) | Not reported | Inclusive curricula |
| Flores 2016 | USA | To reflect (as a teacher) on her experience of using of diverse LGBTQ+ inclusive texts in a primary school | Other (scholarly commentary) | Not reported | Inclusive curricula |
| Francis 2019 | South Africa | To explore how queer youth take up, question and say what they need from sexuality education | Qualitative | LGB secondary school students (n=19) | Inclusive curricula |
| Francis 2019b | South Africa | to explore how counter normative sexualities are discursively constructed in the sexuality education classroom and with what effects by drawing on in-depth interviews with teachers and classroom observation | Qualitative | Secondary school teachers aged 25-63 (n=33) | Inclusive curricula |
| Fulcher 2017 | Australia | To explore heterosexual young people’s perspectives on homophobic language use at school. | Qualitative | Young people aged 16-21 (n=16) | Inclusive anti-bullying and harassment policies |
| Ginicola et al. 2016 | USA | To review the issues involved and a specific framework for school counsellors who wish to set up a Safe Schools Initiative in the context of a resistant atmosphere, using a social justice framework. | Review | Not reported | Inclusive anti-bullying and harassment policies |
| Green et al. 2018 | USA | To investigate how implementation challenges for sexual and gender minority (SGM) guidelines in schools align with, expand on, and contrast with existing knowledge of contextual factors at the inner and outer context of implementation. | Qualitative | School administrators (n=41) and school health professionals, e.g. school nurse, social worker, counsellor (n=55) | Inclusive anti-bullying and harassment policies |
| Hall et al. 2018 | USA | To examine responses from the adults who attended a photovoice exhibit about LGBTQ-related issues in the school context regarding how the event influenced them as well as quality and satisfaction related to the event. | Quantitative | Adults who attended the photovoice exhibit (n=20) | One-off workshops and media Interventions |
| Harris et al. 2021 | England | To explore the experiences of students who identify as LGBT+ in six secondary schools in the south of England. | Qualitative | Students from six schools (n=38) | GSA and similar student clubs; inclusive curricula |
| Hatzenbuehler and Keyes 2013 | USA | To evaluate whether anti-bullying school policies that are inclusive of sexual orientation are associated with a reduced prevalence of suicide attempts among lesbian, gay, and bisexual youths. | Quantitative | 11th-grade public school students (n=31,852) | Inclusive anti-bullying and harassment policies |
| Ioverno et al. 2016 | USA | To examine the influence of presence of and participation in a GSA on perceptions of safety at school, homophobic bullying experiences, and psychosocial adjustment (depression and self-esteem) in 327 LGBTQ students across two school years | Quantitative | LGBQ cisgender students (n=327) | GSA and similar student clubs |
| Ioverno et al. 2021 | Italy | To examine whether students’ observations of teacher and peer interventions against homophobic name-calling and perceptions of the representation of lesbian, gay, bisexual, and transgender (LGBT) issues in class are associated with student intervention behaviours against homophobic name-calling | Quantitative | High school students (n=1,470) | Inclusive curriculum; ally and staff training |
| St John et al. 2014 | Canada | To explore the roles of GSAs in creating supportive school environments for LGBTQ youth and allies. | Qualitative | Youth, teachers and a key informant LGBTQ youth service provider (n=15) | GSA and similar student clubs |
| Jones and Hillier 2012 | Australia | To explore what might be meant by ‘good school sexuality education’ for GLBTIQ students, with a focus on policy-based approaches by focusing on the following research questions: Who are GLBTIQ students and what is their school sexuality education experience? What constitutes ‘good school  sexuality education’ for this group? What are the obstacles to its provision? How can these be overcome? | Mixed methods | Australian GLBTIQ young people (n=3,134) | Inclusive anti-bullying and harassment policies |
| Konishi et al. 2013 | Canada | To examine whether students' odds of recent substance use were lower in the presence of gay-straight alliances or explicit anti-homophobia policy that had been established at their school recently, or at least 3 years prior. | Quantitative | Secondary school students (n=21,708) | GSA and similar student clubs; inclusive anti-bullying and harassment policies |
| Kosciw et al. 2012 | United states | To examine the effects of a negative school climate on achievement and the role that school-based supports—safe school policies, supportive school personnel, and (GSA clubs—may have in offsetting these effects. | Quantitative | LGBT secondary school students (n=5,730) | GSA and similar student clubs; inclusive anti-bullying and harassment policies; inclusive curricula |
| Kroneman et al. 2019 | Netherlands | To examine the effects of an intervention on sexual prejudice in prevocational secondary schools. | Quantitative | 8th-grade students (n=60) | One-off workshops and media interventions |
| Kull et al. 2016 | USA | To examine the relationship between antibullying policies and LGBT students’ safety and victimization at school. | Quantitative | 6-12th grade students (n=8,584) | Inclusive anti-bullying and harassment policies |
| Lucassen and Burford 2015 | Aotearoa/New Zealand | To evaluate the potential of a 60-minute sexuality diversity workshop to address bullying in secondary  schools. | Mixed methods | Secondary students (n=237) | One-off workshops and media interventions |
| Mayberry et al. 2011 | USA | To explore the efficacy of GSAs and identify school practices that either support or destabilize antigay school environments. | Qualitative | GSA student members at high-school level (n=12), GSA advisors (n=4), high school principals (n=2), district administrators (n=2) | GSA and similar student clubs |
| Mitton-Kukner et al. 2016 | Canada | To explore the impact of Positive Space training on pre-service teachers’ understanding of and abilities to create safe spaces for LGBTQ youth and allies in schools in response to the anti-homophobia/transphobia and LGBTQ-inclusive training they received as part of formal teacher training | Qualitative | Pre-service teachers (n=9) | Ally and staff training |
| O'Farrell et al. 2021 | Ireland | To appraise and synthesis the evidence in relation to both the receipt and delivery of LGBTI+ inclusive sexual health education. | Review | NA | Inclusive curricula |
| Ollis 2010 | Australia | To explore impact of the professional development activities on teachers’ attitudes to homosexuality and their ability to address homophobia | Qualitative | Secondary school teachers (n=14) | Ally and staff training |
| Poteat et al. 2019 | Norway | To examine factors that could account for which teachers report (a) more consistently intervening against homophobic language use when they observe it and (b) more frequently discussing homophobic language with their students in their classes. | Quantitative | Teachers (n=283) | Ally and staff training |
| Rabbitte 2020 | USA | To examine sexual health education programmes in schools in the USA for the inclusion of information on gender identity and sexual orientation. | Review | Primary and secondary/high school children (n not reported) | Inclusive curricula |
| Russel et al. 2016 | USA | To examine the role of SOGI-focused policies in association with bullying | Quantitative | NA (Survey data based on school principal and teacher reports and administrative data) | GSA and similar student clubs; inclusive anti-bullying and harassment policies |
| Saewyc et al. 2014 | Canada | To explore the relationship between school-based GSA and explicit anti-homophobic bullying in secondary schools in Canada with experiences of anti-gay discrimination, suicidal ideation and attempts among LGB, mostly heterosexual and exclusively heterosexual students. | Quantitative | 7-12th-grade school students (n=21,708) | GSA and similar student clubs |
| Saewyc et al. 2016 | Canada | To identify and evaluate the existing research about school-based interventions to improve outcomes for LGBTQ youth, examine the quality and relevance of the evidence for schools in Canada, and weigh the potential benefits of different school interventions compared to the potential costs of the health outcomes they address. | Review | 13 studies of interventions and their impacts on health outcomes for LGBTQ+ youth | GSA/similar student clubs; inclusive anti-bullying and harassment policies; inclusive curricula |
| Schijf et al. 2020 | Philippines | To explore the Human Library Program of the DLSU Integrated School Libraries aiming to foster diversity and reduce prejudice and discrimination against people with different backgrounds including members of the LGBTQ community | Mixed methods | 5-12th-grade students (n not reported) | Inclusive curricula |
| Sinacore et al. 2018 | Taiwan | To examine faculty and staff's perception of the implementation of Gender Equity Education Act (GEEA) and its influence on the school community. | Qualitative | Middle and high school teachers aged 28 to 45 (n=15) | Inclusive anti-bullying and harassment policies; ally and staff training |
| Singh et al. 2013 | USA | To explore the experiences of group leaders facilitating popular opinion leader (POL) groups aimed at reducing LGBTQQ aggression in middle school | Qualitative | Group leaders (n=8) and student popular opinion leaders (n=40) | Other and one-off workshops and media interventions |
| Snapp 2015 | USA | To examine whether student's perceptions of personal safety and school climate safety are stronger in the presence of LGBTQ-inclusive curricula | Quantitative | Students (n=1,232) | Inclusive curricula |
| Steck and Perry 2018 | USA | to identify perceptions of experiences creating a safe and inclusive environment for students who identified as LGBTQ. | Qualitative | Secondary school administrators (n=7) | Inclusive anti-bullying and harassment policies |
| Swanson and Gettinger 2016 | USA | To explore teachers outcomes, including knowledge and attitude toward LGBT youth in relation to 3 school-wide policies designed to support LGBT students | Quantitative | 6-12th-grade teachers (n=98) | Ally and staff training |
| Truong and Zongrone 2021 | USA | To explore the psychosocial benefits of GSA participation. | Quantitative | LGBTQ secondary school students (n=11,164) | GSA and similar student clubs |
| Van de Ven 1995 | Australia | To evaluate outcomes of an antihomophobia teaching kit for students in a pre-test post-test follow-up design | Quantitative | High school students aged 13-16 (n=130) | Inclusive anti-bullying and harassment policies |
| Vilkin et al. 2019 | USA | To examine how an afterschool arts-based curriculum for grades K–5 that embraced expansive understandings of gender was related to children’s gender attitudes and beliefs | Quantitative | Students in after-school programmes (n=83) | Inclusive curricula |
| Wernick et al. 2013 | USA | To determine the effectiveness of a programmatic intervention developed and administered by LGBTQ youth that seeks to increase knowledge and awareness about homophobia and transphobia as well as students' likelihood and confidence to intervene when offensive language or actions target LGBTQQ students in schools. | Quantitative | High school students and middle school 8th-graders (n=537) | One-off workshops and media interventions |
| Wernick et al. 2016 | USA | To 1) investigate if participation in an intervention designed and led by LGBTQQ youth using theatre and dialogue is related to increases in participants’ intentions to advocate in support of LGBTQQ communities 2) explore how awareness of homophobia and transphobia is constructed by youth and the relationship between participation in the intervention, homophobia and transphobia awareness, and LGBTQQ advocacy intentions. | Mixed methods | High school and middle school students (n = 515) | One-off workshops and media interventions |

#### Key:

#### GSA: Gay-straight Alliance

#### SOGI: Sexual or Gender Identity

LGBTQQ: lesbian, gay, bisexual, transgender, queer, and questioning

LGBT+: lesbian, gay, bisexual, transgender, queer plus others

### Supplementary Table 3: Context-Mechanism-Outcome (CMO) configurations for Gay-Straight Alliances or similar student clubs

| **Gay Straight Alliances or similar student clubs** | |
| --- | --- |
| **Context, Mechanism, Outcome (CMO)** | **Reference** |
| When schools have safe spaces including youth-led GSAs or similar student clubs for LGBTQ students (C) then teachers might report fewer bullying problems (O) when teachers judged schools to be unsafe (C). No mechanism reported. | Russell et al. 2016 |
| When LGBQ high school and university students attended schools with GSAs (C) then they reported reduced homophobic bullying over time (O) as GSAs may contribute to positive overall school climate (M). | Ioverno et al. 2016 |
| When LGBQ high school and university students attended schools with GSAs and participated in their schools' GSAs (C) then they did not report reduced homophobic bullying over time (O). No mechanism reported. | Ioverno et al. 2016 |
| When LGBQ high school and university students attended schools with GSAs and participated in their schools' GSAs (C) then they did not report subsequent increased psychological wellbeing (well-being included self-esteem and depression) (O). No mechanism reported.. | Ioverno et al. 2016 |
| When LGBQ high school and university students attended schools with GSAs and participated in their schools' GSAs (C) then they reported greater subsequent school safety (O) as GSAs may contribute to positive overall school climate and provide safe space for LGBTQ youth (M). | Ioverno et al. 2016 |
| When a school has a GSA or similar club (c) then anti-LGBT victimisation is decreased (no association with self-esteem or educational outcomes) (O). No mechanism reported. | Kosciw et al. 2012 |
| When students who are more severely victimised based on their sexual orientation have a GSA or similar club in their school (c) then they have fewer missed days of school (O). No mechanism reported. | Kosciw et al. 2012 |
| When schools had longer-established GSAs that had been place for at least 3 years (compared to schools without GSAs) (c) then lesbian and bisexual girls had lower odds of consuming alcohol as well as binge drinking (O). This may be because school strategies reduce homophobia and foster safe and supportive environments as well as school inclusion, and reduce stress-related health risks (M). | Konishi et al. 2013 |
| When schools had longer-established GSAs that had been place for at least 3 years (compared to schools without GSAs) (C) then heterosexual students (boys and girls) had lower odds of binge drinking and heterosexual boys had lower odds of drinking any alcohol (O). This may be because school strategies that reduce homophobia and foster school inclusion are also beneficial for heterosexual students since “many students who are harassed for being thought to be gay, lesbian or bisexual may actually identify as heterosexual” (M). | Konishi et al. 2013 |
| If there are GSAs available to students (C) then improvements have been seen in academic performance (O) because students are more comfortable with their sexuality (M). | Black et al. 2012 |
| If students engage with GSAs (C) then students feel more empowered (O) because they feel like they have direct influences on the climate in their schools (M). | Black et al. 2012 |
| If there are GSAs available to students (C) this creates a more positive climate for sexual diversity in a school (O) because GSAs positively affect personal relationships with other students by decreasing bullying (M). | Black et al. 2012 |
| When GSAs are present (C) LGBTQ students report fewer occasions of homophobic and gender-based bullying (O). No mechanism (M). | Day et al. 2020 |
| When GSAs are present (C) LGBTQ students report increased perceptions of social support from classmates (but not teachers) (O). This may be because LGBTQ students experienced fewer occasions of homophobic and gender-based bullying in schools with GSAs (M). | Day et al. 2020 |
| When GSAs are present (C) trans students were twice as likely to experience homophobic bullying compared to cisgender students (O). No mechanism reported. | Day et al. 2020 |
| When GSAs are present (C) Black or African American LGBT youth were less likely to be bullied for homophobic reasons compared to White LGBT youth (very weak to no evidence of a relationship amongst other ethnicities) (O). No mechanism reported. | Day et al. 2020 |
| When GSAs are present (C) Black or African American LGBT youth reported higher perceptions of support from classmates compared to white LGBT youth (very weak to no evidence of a relationship amongst other ethnicities) (O). This may be because they were less likely to be bullied for homophobic reasons compared to White LGBT youth (M). | Day et al. 2020 |
| Even when GSAs are present (C) trans youth reported less classmate support compared with cis youth (O) because trans youth are at higher risk of experiencing homophobic bullying compared to cis youth regardless of GSA presence (M). | Day et al. 2020 |
| When GSAs and LGBT focused policies were present (C) trans girls reported higher teacher support compared to female assigned at birth students (O) because GSAs improve perceptions of social support (M). | Day et al. 2020 |
| When schools in rural settings have a GSA (C) then LGBT students reported higher scores in victimisation compared to schools without a GSA (O) because schools in rural settings may face numerous barriers to successful implementation (e.g. pervasive homophobic attitudes, lack of resources, and few student and teacher allies) (M). | De Pedro et al. 2018 |
| When schools in rural settings have a GSA (C) then LGBT students reported lower scores of feeling safe at school (O) because schools in rural settings may face numerous barriers to successful implementation (e.g. pervasive homophobic attitudes, lack of resources, and few student and teacher allies) (M). | De Pedro et al. 2018 |
| When schools have an LGBTQ+ support group for students (C), it increases inclusivity and decreases isolation (O) because LGBTQ+ students have a space where they do not get judged, feel comfortable, be themselves and make friends (M). | Harris et al. 2021 |
| When staff who identify as LGBTQ+ also attend student LGBTQ+ support groups (C), this leads to a greater sense of inclusivity (O) because it clearly impacts upon the students positively, by enhancing visibility of role models (M). | Harris et al. 2021 |
| When the wider school climate is a supportive safe place, LGBTQ+ young people are more likely to attend LGBTQ+ support groups and feel safe there. If the environment is not supportive (C), GSAs can be harmful for the mental health and well-being of LGBTQ+ young people (O) because peers may make fun of LGBTQ+ students for attending support groups (homophobic bullying) as the support groups enhance visibility and out young people (M). | Harris et al. 2021 |
| When GSAs are present (C) then there is a decrease in bullying (O) because the school climate has improved (M). | Saewyc et al. 2016 |
| When GSAs are present (C) then there is a decrease in suicidal ideation amongst LGB girls and boys (O) because there is an improvement in school climate (M). | Saewyc et al. 2014 |
| When schools have GSAs (C) this positively impacts the emotional safety of LGBTQI+ students (O) because LGBTQI+ students were supported, could build a sense of community, and developed the confidence necessary to “speak out” against antigay behaviours and attitudes in the larger school environment (M). | Mayberry et al. 2011 |
| When schools only implement GSAs to increase acceptance and inclusivity for LGBTQI+ students (C) they might potentially isolate GSA members from the wider school community and not actually address homophobic bullying (O) as LGBTQI+ issues are not addressed beyond club meetings or discussed in the wider school context (M). | Mayberry et al. 2011 |
| When schools have GSAs (C) students feel supported to speak out against derogatory comments, bullying, and other forms of physical and verbal harassment (O) because GSAs provide a sense of community (M). | Mayberry et al. 2011 |
| When schools have longer-established GSAs and explicit anti-homophobic bullying policies (C), then LGB and mostly heterosexual girls (but not LGB or mostly het boys) and exclusively heterosexual boys (but not girls), are less likely to exhibit suicidal ideation and attempts (O). This is because exclusively heterosexual boys are more likely to experience homophobic bullying compared with exclusively het girls (M). | Saewyc et al. 2014 |
| When LGBTQ students are members of GSAs (C) then they are more likely to benefit not only from the direct GSA support but also, they are more likely to be experience less isolation (O). This is because the GSAs help them connect with other LGBTQ community members, events and resources helping them build a sense of community beyond their school experiences (M). | St John et al. 2014 |
| Among LGBTQ students who have a GSA at their school (C), participating in GSAs was not related to improved psychological well‐being, but participating in GSAs was related to greater school belonging and these associations did not differ by racial/ethnic identity (O). This might be because GSAs may provide some degree of socialization among LGBTQ students and allies, but the potential benefits may depend on the types of activities that their GSA engages in (M). | Truong and Zongrone 2021 |
| When there is inclusive programming including GSAs, materials such as posters visible around the school, and relevant discussions included in school curricula (including classrooms learning, assemblies, and workshops) (C) there is reduced homophobic bullying (O). No mechanism reported. | Espelage et al. 2019b |

### Supplementary Table 4: Context-Mechanism-Outcome (CMO) configurations for Inclusive anti-bullying and harassment policies.

| **Context, Mechanism, Outcome (CMO)** | **Reference** |
| --- | --- |
| When GLBTIQ students report that their school has policy-based protection for GLBTIQ students and against homophobia (C) then they are less likely to think about self-harm, self-harm, experience suicidal ideation, and attempt suicide (O). This might be because students experience less verbal, physical, and other types of homophobic abuse in schools with policy-based protection (M). | Jones and Hillier 2012 |
| When GLBTIQ students report that their school has policy-based protection against homophobia (C) then they are more likely to feel safe at school and more likely to rate their school as supportive (O). This might be because students experience less verbal, physical, and other types of homophobic abuse in schools with policy-based protection (M). | Jones and Hillier 2012 |
| When GLBTIQ students report that their school has policy-based protection against homophobia (C) then they are more likely to feel good about their sexuality (O). This might be because students experience less verbal, physical, and other types of homophobic abuse in schools with policy-based protection (M). | Jones and Hillier 2012 |
| When GLBTIQ students report that their school has policy-based protection against homophobia (C) then they are less likely to experience verbal, physical and other types of homophobic abuse (O). No mechanism reported. | Jones and Hillier 2012 |
| When GLBTIQ students report that their school has policy-based protection against homophobia (C) then they are more likely to receive useful information from that school on homophobia and discrimination, gay and lesbian relationships, gay and lesbian safe sex (O). No mechanism reported.. | Jones and Hillier 2012 |
| When a school has a GSA or similar club (c) then anti-LGBT victimisation is decreased (no association with self-esteem or educational outcomes) (O). No mechanism reported. | Jones and Hillier 2012 |
| When GLBTIQ students report that their school has policy-based protection against homophobia (C) then are less likely to report that the school promoted potentially harmful messages, such as 'gay people should become straight' or 'sex before marriage is wrong' and more likely to report more inclusive and affirming messages such as 'homophobia is wrong, '"males" don't have to be "manly" and "females" don't have to be "girly", 'experimenting with sexualities and pleasure is okay' etc. (O). No mechanism reported. | Jones and Hillier 2012 |
| When schools have comprehensive anti-bullying/harassment policy (C) then LGBT students report higher self-esteem (no association with victimisation, GPA, missed days) (O), because policies create an affirming environment (M). | Kosciw et al. 2012 |
| When schools had anti-bullying policies that been place for at least 3 years (C) then lesbian and bisexual girls had lower odds of drinking alcohol (O). This may be because school strategies address homophobia and foster safe and supportive environments as well as school inclusion and reduce stress-related health risks (M). | Konishi et al. 2013 |
| When schools had anti-bullying policies that been place for at least 3 years (C) then lesbian and bisexual girls had lower odds of generally drinking alcohol (O) as policies aim to reduce homophobia and foster school inclusion and connection (M). | Konishi et al. 2013 |
| When schools had longer-established explicit anti-homophobia policies that been place for at least 3 years (compared to those in school with no policies) (c) then heterosexual students (boys and girls) had lower odds of binge drinking, heterosexual girls had lower odds of drinking alcohol as well as binge drinking (O). This may be because school strategies that reduce homophobia and foster school inclusion are also beneficial for heterosexual students since “many students who are harassed for being thought to be gay, lesbian or bisexual may actually identify as heterosexual” (M). | Konishi et al. 2013 |
| When 16-17 year old students attend schools with LGBTQ+ inclusive anti-bullying policies (not solely general restrictive anti-bullying policies) (C), lesbian and gay (though not bisexual or heterosexual) youths are less likely to attempt suicide (O); this might be because risk factors for mental health problems are different in bisexual youth (M). | Hatzenbuehler and Keyes 2013 |
| When schools implement SOGI policies (C) then fewer LGBT students truant (O) because schools are perceived to be more accepting and inclusive (M). | Day et al. 2019 |
| When schools implement SOGI policies (C) then fewer LGB students experienced homophobic bullying (O). No mechanism reported. | Day et al. 2019 |
| When schools implement SOGI policies (C) then more LGB students reported higher school connectedness (O). No mechanism reported. | Day et al. 2019 |
| When anti-homophobia initiatives in schools address homophobic language within broader conversations about social status, reinforcing popularity and masculinity (C), this leads to reductions in homophobic language and slurs (O) because heterosexual students are more likely to respond and change their behaviour. Heterosexual students often do not see themselves as homophobic and do not respond to teaching about that, but they understand ideas about popularity and masculinity and are therefore more likely to respond (M). | Fulcher 2017 |
| When inclusive policies, such as anti-homophobic bullying policies, are implemented (C) there is a decrease in bullying (O) because there may be a decrease in stigma for those who do not fit stereotypes related to gender behaviour (M). | Saewyc et al. 2016 |
| When inclusive policies, such as anti-homophobic bullying policies, are implemented (C) there is a decrease in suicidal ideation (O) because there may be a decrease in stigma for those who do not fit stereotypes related to gender behaviour (M). | Saewyc et al. 2016 |
| When attempting to implement safe school initiatives, and legislation to lower violence and bullying, with an inclusive LGBTQ+ focus, schools in religious settings can face backlash from parents and the community and the intervention can be prohibited (C), which prevents a reduction in bullying and increase in inclusivity (O) because many religious beliefs do not affirm sexual and gender minorities and actively oppose equal rights legislation (M). | Ginicola et al. 2016 |
| When staff are unaware of existing school policies on inclusive anti-bullying (which evidence suggests is a common occurrence) (C) this leads to no change in bullying or inclusivity for sexual and gender minority students (O) because staff do not implement the interventions (M). | Green et al. 2018 |
| When an anti-homophobia teaching kit (a module of six lessons for presentation of a unit on homophobia) is implemented in schools (C), then a significant impact on cognition, homophobic anger, and behavioural intentions might be observed for all students in the short term, but in the long-term effects might be sustained only for homophobic anger and behavioural intentions (O). The intervention's impact on boys’ homophobic cognition was more likely to have attenuated within 3 months (C). This might be because of boy’s socialisation with male homophobia being linked to fears of femininity or lack of manliness (M). There might be scope in targeting sex-role expectations when aiming to reduce homophobia and/or provide follow-up activities for male students. | Van de Ven 1995 |
| When an anti-homophobia teaching kit is implemented in schools (C), then it might not successfully reduce all aspects of homophobia, such as homophobic guilt (both co-educational and single school students) and affect (homophobic guilt, homophobic anger, and delight) for coeducational students) (O). No mechanism reported. | Van de Ven 1995 |
| When districts had antibullying policies based upon sexual orientation, gender identity, and/or gender expression (SOGIE-inclusive policies) (C) then LGBT students reported greater school safety, less victimisation (physical and verbal harassment) based on their sexual orientation and gender expression, and less social aggression (deliberate exclusion, electronic harassment, lies/mean rumours) compared to students with generic policies or no/unidentified policies (O). This is because specific SOGIE-inclusive policies affect institutional culture, including behaviours (less bullying and more intervening), attitudes, and awareness (M). | Kull et al. 2016 |
| If schools have policies and practices focused on sexual orientation and gender identity (SOGI) (C), then reports of bullying might be reduced (O). This may be because SOGI policies create a climate of safety that is related to less bullying (M). Adopting multiple SOGI-focused programs and practices may be most beneficial to schools that are least safe, or where they are needed most. | Russell et al. 2016 |
| The presence of multiple SOGI-focused policies (C) might have stronger influence on bullying problems than any single policy on its own (O). No mechanism reported. | Russell et al. 2016 |
| When school administrators try to create a safe and inclusive school environment for the LGBTQ student population in their schools (C), then they might face barriers in confronting and deconstructing heteronormativity (O). This might be because challenging heteronormativity to achieve transformative change is highly dependent on changing belief systems and ways of behaving toward those who identify as LGBTQ (M). | Steck and Perry 2018 |
| If there is a school-wide approach to uniform, allowing self-determination,students are able to choose clothes they feel represent their gender expression (C), this leads to increased inclusivity (e.g in physical activity) and improved well-being (O), as itsupports and empowers trans and gender diverse students and allows cisgender students to choose too (e.g., girls can wear trousers) (M). | Evans and Rawlings 2021 |
| If schools have anti-discrimination policies/safe schools' policy (C), then fewer suicide attempts are reported (O) because there is an increase in positive school climate (M). | Black et al. 2012 |
| If teachers adopt a social justice framework on LGBTQ issues (C), then students feel more accepted and belongingness in school (O), because there is an increased sense of comfortability to discuss these issues (M). | Bellini 2012 |
| If emotional safety is not incorporated into Safe Schools policy (C), then LGB students may succumb to mental health distress (O) because they do not feel safe or supported by schools (M). | Bellini 2012 |
| When the school leadership is not supportive of gender equity policies and holds patriarchal values and heteronormativity (C) then gender equity acts might not be implemented and when discussed the focus is on heterosexual relationships (O). This is because there might be a systemic lack of attention to sexuality and gender diversion in the programmes as a whole (M). As a result, faculty and SMG students might perceive climate as unsafe and become silenced and marginalised (O). | Sinacore et al. 2018 |
| When there are explicitly inclusive official school policies which include any language that is part of the published, governing, code of the school that asserts intolerance of discrimination or violence on the basis of sexuality or gender status or perceived sexuality or gender status (C) there is decreased homophobic aggression (O). No mechanism reported.. | Espelage et al. 2019b |
| When gender equity education is implemented in schools hostile to LGBT individuals (C), then SMG might be punished, isolated or bullied to their gender expression (O). This might be because holding negative attitudes towards gender equity education might influence social interactions (M) | Sinacore et al. 2018 |
| When teachers have supportive practices (C) then fewer LGBT students experienced homophobic bullying (O). No mechanism reported. | Day et al. 2016 |
| When teachers have supportive practices (C) then more LGBT students reported higher school connectedness (O). No mechanism reported. | Day et al. 2016 |

### Supplementary Table 5: Context-Mechanism-Outcome (CMO) configurations for Inclusive curricula

| **Context, Mechanism, Outcome (CMO)** | **Reference** |
| --- | --- |
| When schools have more extensive sexuality education at the beginning of the school year covering different topics including sexual orientation and gender, resources, STI prevention, relationships and anatomy (C) then students perceived their teachers or school personnel to be more willing to intervene when witnessing LGBTQ name-calling; female students (but not male students) perceived fellow students to be more willing to intervene; and male students (but not female students) reported being more likely to intervene themselves at the end of the school year (O). No mechanism reported. | Baams et al. 2017 |
| When schools have more extensive sexuality education at the beginning of the school year covering different topics including sexual orientation and gender, resources, STI prevention, relationships and anatomy (C) then female (but not male) students perceived LGBTQ name-calling to be reduced at the end of the school year (O). No mechanism reported. | Baams et al. 2017 |
| When students perceive the representation of LGBT issues in class as positive (C) they were more likely to intervene against HNC and to observe other classmates intervene (O). This is because a positive representation of LGBT issues may offer an opportunity to understand the experiences related to the different sexual and gender identities and thus reduce the tolerance for prejudicial attitudes and raise awareness about the seriousness of HNC (M). | Ioverno et al. 2021 |
| When school have inclusive curricula teaching about positive representation of LGBT people, history, and events (C) then LGBT students report less victimisation and higher academic achievement (no association with self-esteem or missed days of school) (O), as inclusive curricula promote respect and equity for all (M). | Kosciw et al. 2012 |
| When secondary school students were exposed to inclusive sexual health education (C), there was less bullying of SGM youth (O), because the existence of these students was normalised (M). | Rabbitte 2020 |
| When Out in Schools events are hosted (C) then lesbian and bisexual girls report fewer instances of homophobic discrimination (O) as the intervention improves school climate (M). | Burk et al. 2018 |
| When Out in Schools events are hosted (C)then lesbian and bisexual girls report lower odds of verbal harassment and social exclusion (O) because the intervention improves school climate (M). | Burk et al. 2018 |
| When Out in Schools events are hosted (C), then gay and bisexual boys report lower odds of verbal bullying (especially teasing) (O) as the intervention improves school climate (M). | Burk et al. 2018 |
| When inclusive curriculums are implemented (C) there is a decrease in bullying (O) as there is an improvement in school climate (M). | Saewyc et al. 2016 |
| If primary school students are exposed to LGBTQ+ literature in a meaningful way that does not feel tacked on (C) this increases inclusivity and acceptance (O) as they learn that there are lots of people who are different, many people who are different have done great things (e.g. Harvey Milk) and it is okay to be different (M). | Flores 2016 |
| When students are taught about LGBT sexuality in school in an open and honest way and teachers are comfortable, with a good sense of humour, and ask directly whether students have questions (C) this makes students feel included and accepted in school (O) as students that being gay is normal and not a bad thing and that they are affirmed and supported (M). | Francis 2019 |
| When the curriculum includes diversity e.g ensuring workbooks accurately include and educate on LGBTQ+ issues (C), this improves inclusivity and acceptance of LGBTQ+ students (O) as they feel interested and engaged and as though their needs and concerns are included and important (M). | Evans and Rawlings 2021 |
| If Out in Schools events are hosted (C) then LGB students report higher feelings of school connectedness/belonging (O) as the intervention improves school climate (M). | Burk et al. 2018 |
| When LGBTQ+ content is included in curricula at a younger age (rather than only for 16+) (C), this increases students’ acceptance of themselves (O) as students feel normalised at an age when they are more likely to be developing their sexual orientation (M). | Harris et al. 2021 |
| When human library projects are integrated in the school curriculum (C) then student readers are more likely to have reduced prejudice against people with diverse backgrounds (O). This might be because the program raises awareness and promotes dialogue, diversity, and empathy among student readers (M). | Schijf et al. 2020 |
| Curricular inclusions of diversity (C) promote acceptance and support for their transgender and gender diverse peers (O) as they educate cisgender students within the class about gender diversity (M). | Evans and Rawlings 2021 |
| When the curriculum includes diversity e.g ensuring workbooks accurately include and educate on LGBTQ+ issues (C), this improves inclusivity and acceptance (O) as it normalises being LGBTQ and leads to people learning and asking questions (M). | Evans and Rawlings 2021 |
| When gender-focused arts-based curricula are offered to children in kindergarden (age xx) through to 5th grade (age xx) (C), then children might gain increased awareness of gender norms, shifts in understandings of gender, and more positive attitudes toward gender-expansive roles, activities, and attire (O). No mechanism reported. | Vilkin et al. 2019 |
| When students who are more severely victimised based on their gender-expression and/or are in schools with poor climates have inclusive curricula teaching about positive representation of LGBT people, history, and events in schools with poor climate in schools (C) then this positively influences the self-esteem of these students (O). No mechanism reported. | Kosciw et al. 2012 |
| When curricular inclusions of diversity avoid overly focusing on ‘deficit and ‘at risk’ narratives (C) this makes students more likely to accept themselves and others (O) as it creates the sense that there is less to be afraid of when being LGBTQ and it is not a negative characteristic (M). | Evans and Rawlings 2021 |
| If students attend schools with LGBTQ+ inclusive curricula (C), the mental health and well-being of LGBTQ+ students improves (O) as LGBTQ+ students feel validated and role models who make valued and important contributions are visible and normalised (M). | Fleshman 2019 |
| If *Out in Schools* events are hosted (C) then lesbian and bisexual girls report lower odds of suicidal ideation because the intervention improves school climate (O). No mechanism reported. | Burk et al. 2018 |
| If *Out in Schools* events are hosted (C) then heterosexual girls report lower odds of suicidal ideation because the intervention improves school climate (O). No mechanism reported. | Burk et al. 2018 |
| A curriculum that does not assume compulsory heterosexuality (C) creates an inclusive learning environment (O) as it shows students that the power of heterosexuality and its associated prejudice and discrimination is not endorsed within that school community (M). | Francis 2019 |
| When sexual minorities are presented positively in teaching in terms of family, stability, love and commitment (C), this creates an inclusive school climate that is not seen as regulating sexuality (O) as students see sexual minorities as equal to heterosexuals rather than seeing them through a lens of deviance, deficit and risk (M). | Francis 2019b |
| An inclusive and comprehensive sexual education curriculum (C) has been associated with decreased homophobic bullying (O). Nomechanism reported. | Espelage et al. 2019b |
| When LGBTI+ topics were included in sexual health education, such as same sex relationships, LGBTI+ terminology, sexual orientation, gender identity, approaches for STI and HIV (C) this reduced stigma, fostered self-esteem and limited negative mental health outcomes for LGBTQI youth (O). No mechanism reported. | O’Farrell et al. 2021 |
| When LGBTI+ topics were included in sexual health education (C) then LGBTI+ young people showed increased well-being (O) as they saw themselves represented in the curriculum (M). | O’Farrell et al. 2021 |
| When teachers lack the competency and knowledge to deliver existing gender equity education (C) then SMG students might be marginalised and isolated when discussing gender issues in the classroom (O) as teachers might predominantly focus on heterosexual intercourse with little or no attention to sexual and gender diversity due to inadequate training and poor coordination (M). | Sinacore et al. 2018 |
| The second step programme - a social emotional learning intervention (C) did not lead to reduction in bullying, homophobic name calling, cyberbullying or sexual harassment (O). No mechanisms reported. | Espelage et al. 2019 |
| At the school level, when schools have LGBTQ-inclusive curricula especially in sexuality education/health classes (C), then it is likely that there is more school safety and less bullying than schools with less supportive curricula (O). This might be because when lessons are viewed as more supportive safety increases and bullying decreases (M). | Snapp 2015 |
| At the individual level, when students receive LGBTQ-inclusive curricula (C), then they might be more likely to report perceptions of safety compared to same school students that did not receive these curricula (O). This might be because inclusive curricula improve the overall school climate (M). | Snapp 2015s |
| At the individual level, when students receive LGBTQ-inclusive curricula (C), then they might be more likely to report being bullied compared to same school students that did not receive these curricula especially when supportive lessons were taught in mathematics/science, music/art/drama and PR courses (O). This might be because the presence of inclusive curricula may heighten students’ awareness of bullying and safety, or schools may teach inclusive curricula in schools where the climate for LGBTQ youth is already unsafe (M). | Snapp 2015 |
| When schools have supportive LGBTQ-inclusive curricula (C), then reports of bullying might increase at the individual level but the overall positive effects of these strategies on school climate may outweigh any negative associations (O). No mechanism reported. | Snapp 2015 |

### Supplementary Table 6: Context-Mechanism-Outcome (CMO) configurations for workshops including media interventions

| **Context, Mechanism, Outcome (CMO)** | **Reference** |
| --- | --- |
| When gender-diverse/transgender people share their stories with secondary school students during a 60-min gender diversity workshop to address bullying and promote positive environments for learning (C) then one student perceived this as especially helpful for understanding gender/diverse people (O) as students “kn(e)w how it feels to be trans/gay” (M). | Burford et al. 2017 |
| When secondary students attended a 60-min gender diversity workshop to address bullying and promote positive environments for learning (C) then students valued and understood gender/diverse people significantly more and had more accepting views after compared to prior to the workshop (O) as the workshop raised awareness about transgender discrimination and increased empathy towards transgender people (M). | Burford et al. 2017 |
| When secondary students attended a 60-min gender diversity workshop to address bullying and promote positive environments for learning (C) then 80% of students thought that the workshop would reduce bullying in schools (O) as the workshop raised awareness about transgender discrimination and increased empathy towards transgender people (M). | Burford et al. 2017 |
| When secondary students attended a 60-min sexuality diversity workshop which included a person with lived experiences discussing their “coming out” experience to address bullying in schools (C) students self-reported more accepting and supportive attitudes towards sexuality diverse individuals after compared to prior to the workshop (O) as the workshop, especially the person with personal experiences, increased students’ empathy towards sexuality diverse individuals as well as increased their understanding of experiences of homophobia and bullying (M). | Lucassen and Burford 2015 |
| When secondary students attended a 60-min sexuality diversity workshop which included a person with personal experience discussing their “coming out” experience to address bullying in schools (C) 75.8% of students thought the workshop will reduce bullying in schools (O) as the workshop, especially the person with personal experiences, increased students’ empathy towards sexuality diverse individuals as well as increased their understanding of experiences of homophobia and bullying (M). | Lucassen and Burford 2015 |
| When Colour of the Rainbow workshops were given (C) then students reported less homophobic bullying (O) because they were able to explore their own attitudes and subsequently change them (M). | Douglas et al. 2010 |
| When Colour of the Rainbow workshops was given (C) then students reported less homophobic bullying (O) because they were able to explore their own attitudes and subsequently change them (M). | Douglas et al. 2010 |
| When LGBT Speaker Panels are conducted (C) then students report being more tolerant of LGBT people (O) because personal story activities appear to promote tolerance and acceptance (M). | Eick et al. 2016 |
| When students received a peer intervention to promote respect for LGB students (C), students showed less positive attitudes towards gender and sexual diversity after the intervention (O). No mechanism reported. | Kroneman et al. 2019 |
| When students received a peer intervention to promote respect for LGB students (C), male students showed a lower intention to help a bullied classmate, but female students should an increased intention to help a bullied classmate after compared to prior to the intervention (O). No mechanism reported. | Kroneman et al. 2019 |
| When students received a peer intervention to promote respect for LGB students (C), students were less positive towards lesbians and gay men, and towards gay and bisexual classmates after the intervention (O). No mechanism reported. | Kroneman et al. 2019 |
| When students received a peer intervention to promote respect for LGB students (C), students were less positive towards lesbians and gay men, and towards gay and bisexual classmates after the intervention (O). No mechanism reported. | Kroneman et al. 2019 |
| When students received a peer intervention to promote respect for LGB students (C), students were more positive about the possibility of coming out at school after the intervention (O). No mechanism reported. | Kroneman et al. 2019 |
| When students received a peer intervention to promote respect for LGB students (C), female students were more positive towards had more positive attitudes towards gay and bisexual classmates compared to male students (O). No mechanism reported. | Kroneman et al. 2019 |
| When students received a peer intervention to promote respect for LGB students (C), female students had more positive attitudes towards lesbians and gay men after compared to prior to the intervention, but male students’ attitudes didn’t change (O). No mechanism reported. | Kroneman et al. 2019 |
| When students received a peer intervention to promote respect for LGB students (C), female students showed more positive attitudes towards gay and bisexual male students and lesbian and bisexual female students when compared to male students (O). No mechanism reported. | Kroneman et al. 2019 |
| When a photovoice intervention by LGBTQ+ school students was held in a small rural town and attended by adults from the community (C), 81% of adults stated they planned to take action or behave differently as a result of attending the intervention. The most common theme involved adults being more supportive or affirming of LGBTQ youths moving forward (O), because the intervention had raised awareness about the oppression these young people experienced and generated feelings of empathy in the adults - taking the perspective of the LGBTQ students to understand and emotionally connect with the issues they were facing (M). | Hall et al. 2018 |
| When students viewed a theatre performance and participated in a post-performance dialogue on the topic of anti LGBTQQ+ bulling through the sharing of LGBTQQA students' lived experiences, identities and personal narratives (C), this increased their likelihood to intervene and confidence to successfully intervene when witnessing anti-LGBTQQ bullying (O) as they learned concrete information and skills to support their ally behaviours regarding bullying and harassment of sexual and gender minority youth (M). | Wernick et al. 2013 |
| When students viewed a scripted 35–40-minute theatre performance on the topic of experiences of heterosexism and genderism and participated in a post-performance 'common ground' exercise and small group discussion on the topic of issues related to identity, LGBTQQ communities, and bullying and harassment (C), this increased reports of willingness to advocate for LGBTQ+ students, and built awareness about homophobia and transphobia (O). This might be because the performance influenced the students to rethink their assessment of whether or not homophobia/transphobia was a problem in their school, and bolstered their considerations of the severity of homophobia/transphobia (M). | Wernick et al. 2016 |
| The use of Popular Opinion Leader (POL) Groups in middle schools (C) might reduce homophobic bullying (O). This might be because behavioural norms may be changed if 15% of the school cohort shifts their behavioral norms to more positives ones, as this shift will diffuse throughout the entire cohort over time (M). | Singh et al. 2013 |

### Supplementary Table 7: Context-Mechanism-Outcome (CMO) configurations for LGBTQ+ ally and staff training

| **Context, Mechanism, Outcome (CMO)** | **Reference** |
| --- | --- |
| When teachers and school staff are well-informed on LGBTQ+ and gender issues, they are more likely to initiate the creation of safe spaces (C), which improves their well-being (O), as LGBTQ+ students can relax and de-stress in said safe spaces (M). | Evans and Rawlings 2021 |
| If teachers and school staff are well-informed on LGBTQ+ and gender issues, they are more likely to refer LGBTQ+ students to appropriate sources of support in the community, counselling or psychological services (C), which improves mental health and well-being (O) as students can build connections, feel accepted and receive treatment (M). | Evans and Rawlings 2021 |
| Teachers and school staff who are well-informed about LGBTQ+ and gender issues are more likely to use the correct pronouns and names for trans and gender diverse students (C) which improves mental health and well-being (O). This was because students report: “I think, it was really weighing me down, because as soon as it had happened I was like ahh I can focus on my studies now because I have one less thing to think about, nobody’s misgendering me, nobody’s calling me the wrong name. Everybody knows what’s up. It’s cool” (M). | Evans and Rawlings 2021 |
| Teachers and school staff who are well-informed about LGBTQ+ and gender issues are more likely to respect the confidentiality of students, for example by not sharing that the student is trans, gender diverse, non-heterosexual, without their consent (C), which improves mental health and well-being (O) as it protects the student against negative treatment and makes them feel safe and protected (M). | Evans and Rawlings 2021 |
| When LGBT students report the school to have a greater number of educators who are supportive of LGBT students (C) then they experienced less victimisation, greater self-esteem, higher GPAs, and fewer missed days of school (O). This might be because supportive staff might provide a personal connection helping keep students in school and buffering against severe victimisation. Staff might also create safe and affirming environments by intervening during homophobic remarks and victimisation, providing support for individual students and advocating for school-wide efforts, such as affirming and protective policies and practices among staff and administration (M). | Kosciw et al. 2012 |
| When teachers in training received Positive Space training (C) this helped them to acknowledge and address the gender binary in school (O), as the training made them more aware of the gender binary and its presence in schools (M). | Mitton-Kukner et al. 2016 |
| When teachers in training who lack experience with the LGBTQ community and do not have GSAs at their schools received Positive Space training (C) this helped them better understand the inclusive nature and purpose of GSAs (O). No mechanism reported. | Mitton-Kukner et al. 2016 |
| When teachers want to support LGBT students then the most frequent barriers (C), they might face are limited training and resources (i.e., lack of training, knowledge, time, and LGBT-inclusive curriculum) (O). This might be because LGBT training may increase teachers’ awareness of the challenges faced by LGBT students and pinpoint ways to provide support, thereby promoting engagement with LGBT students and engendering positive attitudes (M). | Swanson and Gettinger 2016 |
| When teachers attend a sexuality workshop (C) this changed teachers’ personal attitudes and positioning to issues of sexuality diversity (O) as they were able to place themselves in someone else’s mindset (M). | Ollis 2010 |
| If teachers and school staff who are well-informed about LGBTQ+ and gender issues: For example, the school librarian introduced a sticker system to identify books with LGBT themes and/or characters (C), which improves mental health and well-being (O) as students were more likely to perceive the school and learning environment as safe, accepting and progressive (M). | Evans and Rawlings 2021 |
| If teachers and school staff who are well-informed about LGBTQ+ and gender issues: For example, the school librarian began lunch time film screenings with diversity in the characters represented and many LGBT themed films (C), which improves mental health and well-being (O) as students were more likely to perceive the school and learning environment as safe, accepting and progressive (M). | Evans and Rawlings 2021 |
| When teachers in training who felt uncomfortable talking about the LGBTQ community and LGBTQ issues received Positive Space training (C) this helped them to address homophobia and transphobia in class (O), as the training provide them with the necessary language to intervene and discuss LGBTQ issues (M). | Mitton-Kukner et al. 2016 |
| When teachers in training received Positive Space training (C) this increased their awareness and comfort discussing and intervening when witnessing a homophobic or transphobic act (O), as the training showed teachers how to react and proactively create positive spaces for LGBTQ youth (M). | Mitton-Kukner et al. 2016 |
| When teachers attend a sexuality workshop (C) this impacted teachers’ awareness and ability to respond to homophobia (O) as they were better equipped to deal with homophobia (M). | Ollis 2010 |
| When teachers received professional development related to homophobic teasing, preventing students from engaging in name-calling using homophobic slurs, an addressing students’ engagement in name-calling using homophobic slurs (C) this was associated with discussing homophobic language use with students in their class (O). This might be because professional development courses may give teachers the language to express their general disapproval of homophobic behaviour and information to share with students about discrimination against LGBTQI+ youth (M). | Poteat et al. 2019 |
| When teachers received professional development related to homophobic teasing, preventing students from engaging in name-calling using homophobic slurs, an addressing students’ engagement in name-calling using homophobic slurs (C) this was not associated with intervening more consistently when students used homophobic language (O). This might be because professional development courses may not adequately prepare teachers to intervene directly when such behaviour occurs (M). | Poteat et al. 2019 |
| When pre-service teachers received Positive Space training (C) this increased their awareness about the LGBTQ community and understanding about the potential challenges that some LGBTQ individuals may experience (O), as the training helped to recognise the severity and impact of homophobia and transphobia (M). | Mitton-Kukner et al. 2016 |
| When teachers work in a school with an active GSA or enumerated antibullying policy and received training specifically related to LGBT youth (C), then they might report a higher frequency of engaging behaviours to support LGBT students (O). No mechanism reported. | Swanson and Gettinger 2016 |
| When teachers intervene (C) when they observe slurs or bullying then LGBT students felt safer and less victimised (O). No mechanism reported. | De Pedro et al. 2018 |
| When peers intervene when they observe slurs or bullying (C) then LGBT students felt safer and less victimised (O). No mechanism reported. | De Pedro et al. 2018 |
| When students observe teachers intervene during episodes of homophobic name-calling (C), then they were more likely to intervene against HNC and to observe other classmates intervene (O) as the teachers’ intervention may communicate clear expectations that HNC behaviours are unacceptable in school (M). | Ioverno et al. 2021 |
| When participants observe other students intervene against HNC (C) they were more likely to intervene themselves (O). This might be because of peer influence and/or modelling the classroom norms (M). | Ioverno et al. 2021 |
| LGBTQ+ students (C) were more likely to alert teachers during episodes of HNC than heterosexual students (O). This may be because LGBTQ+ students might be more motivated to intervene as they are more likely to perceive the seriousness of HNC and to be aware of its negative impact and thus may feel greater responsibility for providing help and be more aware of how to intervene (M). | Ioverno et al. 2021 |
